# GWAS highlights the neuronal contribution to multiple sclerosis susceptibility

**DOI:** 10.1101/2024.12.04.24318500

**Authors:** Lu Zeng, Khan Atlas, Tsering Lama, the International Multiple Sclerosis Genetics Consortium, Tanuja Chitnis, Howard Weiner, Gao Wang, Masashi Fujita, Frauke Zipp, Mariko Taga, Krzysztof Kiryluk, Philip L. De Jager

## Abstract

Multiple Sclerosis (MS) is a chronic inflammatory and neurodegenerative disease affecting the brain and spinal cord. Genetic studies have identified many risk loci, that were thought to primarily impact immune cells and microglia. Here, we performed a multi-ancestry genome-wide association study with 20,831 MS and 729,220 control participants, identifying 236 susceptibility variants outside the Major Histocompatibility Complex, including four novel loci. We derived a polygenic score for MS and, optimized for European ancestry, it is informative for African-American and Latino participants. Integrating single-cell data from blood and brain tissue, we identified 76 genes affected by MS risk variants. Notably, while T cells showed the strongest enrichment, inhibitory neurons emerged as a key cell type, highlighting the importance of neuronal and glial dysfunction in MS susceptibility.

## Introduction

The genetic architecture of multiple sclerosis (MS) has come into focus over the past decade. Efforts have been most successful around genetic susceptibility, with over 233 independent risk variants identified to date (*1*), but a recent study reported one genome-wide significant severity locus (*2*). While the functional consequences of some susceptibility variants have been characterized – such as the protective effect rs2300747^G^ (*3, 4*) within the *CD58* locus and the risk allele rs1800693-G in *TNFRSF1A* (*5*) - most of these variants remain poorly understood, and there have been few dedicated efforts to systematically map such effects (*1, 6–10*). Functional consequences of MS variants have been found primarily in peripheral immune cells and in microglia, the resident mesoderm-derived immune cell in the central nervous system (*1*). While some effects have been noted in non-immune cells, such as astrocytes, in targeted analyses (*11–13*), such studies highlight an important challenge in functional genomics as the effect of risk variants can be seen in multiple different cell types and subtypes, creating ambiguity about which cell type is the causal one or whether a combination of cell types is required. Further, the limited availability of quantitative trait locus mapping results in a cell-type specific manner outside of peripheral blood mononuclear cell (PBMC) populations means that the extent of a variant’s effect beyond PBMC is largely unknown.

Thus, despite some suggestions (*14–16*), there is currently a dearth of evidence that neuroectodermal derivatives that make up the central nervous system are involved in the onset of MS. Rather, the predominance of an initial peripheral auto-inflammatory response is further supported by the fact that approximately half of MS susceptibility variants may be shared with one or more autoimmune disease (*17*); it appears that an important component of genetic susceptibility to MS involves dysregulated pathways that lead to a propensity for auto-reactive immune responses. Interestingly, among the shared loci, a large proportion have an opposite effect in other diseases (an MS risk allele is protective for another disease), and MS shares more susceptibility loci with certain auto-inflammatory diseases, including ulcerative colitis (UC), celiac disease (CeD), inflammatory bowel disease (IBD), psoriasis (PS), and rheumatoid arthritis (RA) than others (*18*). While this portion of shared genetic susceptibility may be more readily understood functionally, the functional consequences of the other MS-specific half of susceptibility variants remains to be determined; it presumably contributes to the targeting of the auto-inflammatory process to the central nervous system instead of the skin, pancreas, joints, or other tissue.

Here, we focused on systematically exploring the question of possible MS susceptibility variants exerting functional consequences only in neuronal and glial cell types. To properly power such a systematic evaluation genome-wide, we accessed our prior MS susceptibility results, expanding discovery meta-analysis with three new genome-wide datasets: the UK Biobank (UKBB) (*19*), the Electronic Medical Records and Genomics (eMERGE) (*20*) study, and the initial release of the All of US cohort (AoU) (*21*). Our team has previously harmonized these three datasets (*22*) into a coherent dataset of 750,051 participants. This significantly expanded the GWAS as our prior study had only a targeted replication effort (*1*). Further, these three cohorts have substantial numbers of diverse participants, allowing us to complete a multi-ancestry meta-analysis in MS and to pose some important questions about the relevance of MS susceptibility loci discovered among participants of European ancestry (EUR), African-American (AFR) and Admixed American (AMR). To identify potential causal MS genes, we integrated the extended EUR GWAS results with the gene expression data from EUR participants using a colocalization analysis (COLOC) across the six major cell types of the dorsolateral prefrontal cortex (DLPFC) and 14 major cell types of the peripheral blood mononuclear cell (PBMC). We then compared the effect of MS risk loci across 12 inflammatory diseases, four neurodegenerative diseases, four psychiatric disorders, and metabolic traits. Finally, we designed, optimized, and tested a genome-wide polygenic score (GPS) (*23*) for MS that maximizes performance across ancestries. We then conducted a hypothesis-free phenome-wide association study (PheWAS) to identify diseases/traits associated with the GPS, and examined the GPS associations with brain MRI data collected from MS patients (**Fig. 1**).

**Fig. 1.**
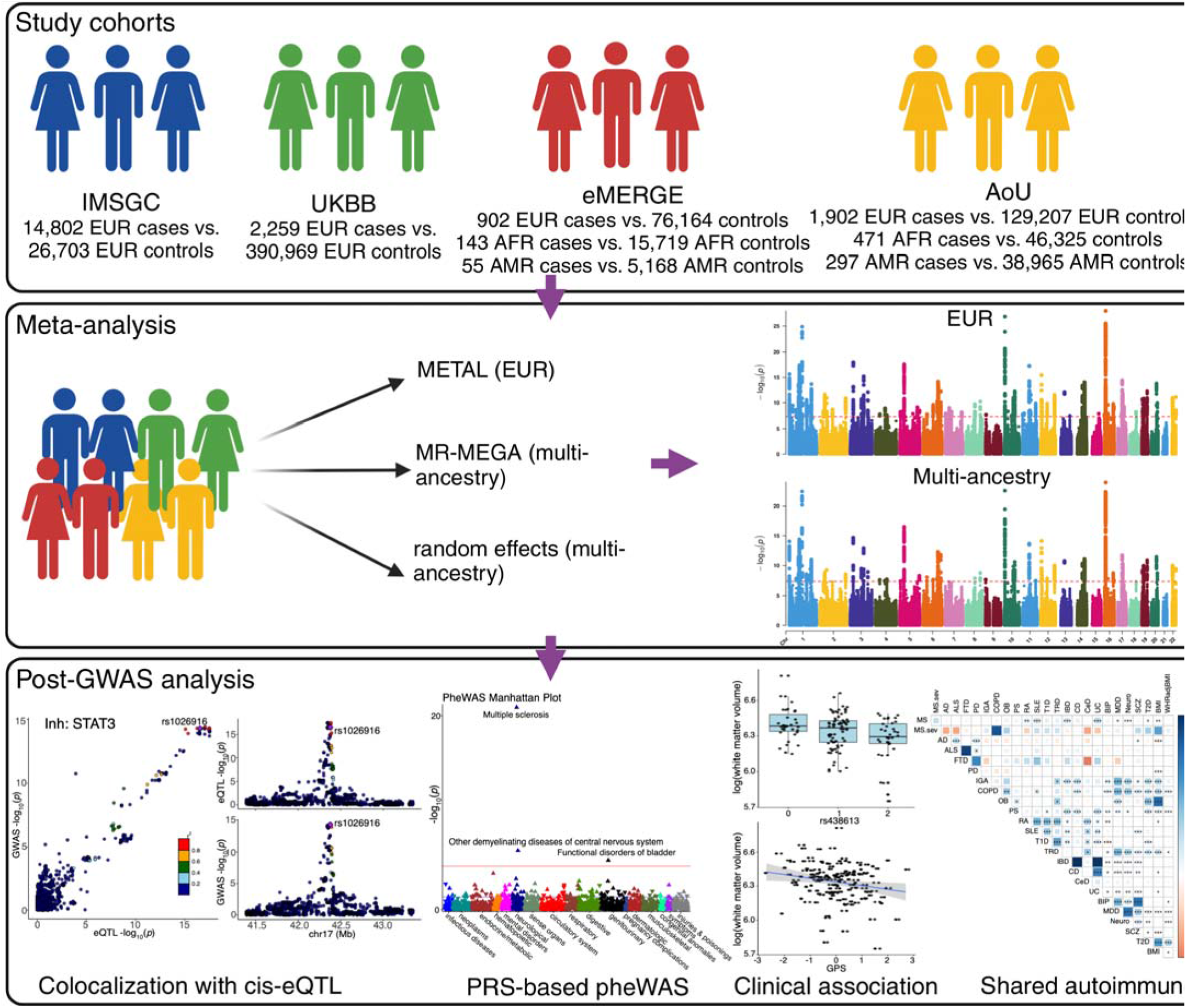
MS GWAS study design. Top panel: four cohorts used in the meta-analysis. Middle panel: meta-analysis and the three methods used. METAL provides a computationally efficient tool for meta-analysis of genome-wide association scans in European ancestry, MR-MEGA (middle) can identify risk variants with heterogeneous effects due to population stratification introduced by ancestry differences, whereas random-effect (bottom) is better suited for risk variants with homogeneous effect direction across different ancestries. The red dashed lines indicate p-value threshold of P < 5 × 10^−8^. Bottom panel: downstream analyses and their examples.

## Results

### New European ancestry GWAS meta-analysis for MS

We first harmonized the genetic and phenotypic data available from the UKBB, eMERGE-III, and AoU datasets (*22*), defining cases by ICD 9: 340, 323 and 341 (**Supplementary table S1**). We then conducted a European ancestry GWAS meta-analysis (using METAL) (*24*) that includes a total of 5,063 MS cases and 596,340 controls (see Methods) (**Supplementary table S1**). These results were subsequently combined with our prior meta-analysis (*1*), increasing sample size to a total of 19,865 MS patients and 623,043 controls. Since the focus of this project was the evaluation of non-immune SNPs, we elected to exclude the extended Major Histocompatibility Complex (MHC) region from our analysis (Chr6: 25,383,722-33,368,421bp in GRCh37).

A total of 5,041 non-MHC SNPs exceeded a threshold of p<5x10^-8^ in the new meta-analysis (**Fig. 2**); 99% of these SNPs showed a concordant direction of effect between the prior study (*1*) and the three new cohorts. Using linkage disequilibrium (LD)-based clumping methods, we identified 236 SNPs independently associated with MS susceptibility among the 5,041 significant SNPs. We then removed SNPs with r^2^> 0.1 and within ±500kb window of any of the 200 previously reported susceptibility variants (*1*). A total of 38 SNPs were not in LD with the previously reported SNPs. Next, we defined novel MS genomic risk loci using non-overlapping genomic segments that contain at least one MS SNP, with the condition that MS SNPs in adjacent loci are more than 250 kb away from each other (that is, a 250-kb window on each side of one of the SNPs). This approach results in 4 loci that do not appear to have been reported previously as harboring MS susceptibility variants (**Table 1**). Therefore, most of the new independently associated variants (n=34) fall within loci that harbor other MS susceptibility variants. We have also removed two susceptibility SNPs reported in our previous study (rs6498163 and rs11256593) due to an LD > 0.1 with other MS SNPs; in these cases, we kept the SNP within a pair that had the smaller p-value, which brings the total count of current MS susceptibility variants to 198 known and 38 novel variants, or 236 independent MS susceptibility effects, each of which is labeled by a lead SNP (**Supplementary table S2**). We used the results of this new meta-analysis for all subsequent analyses.

**Fig. 2.**
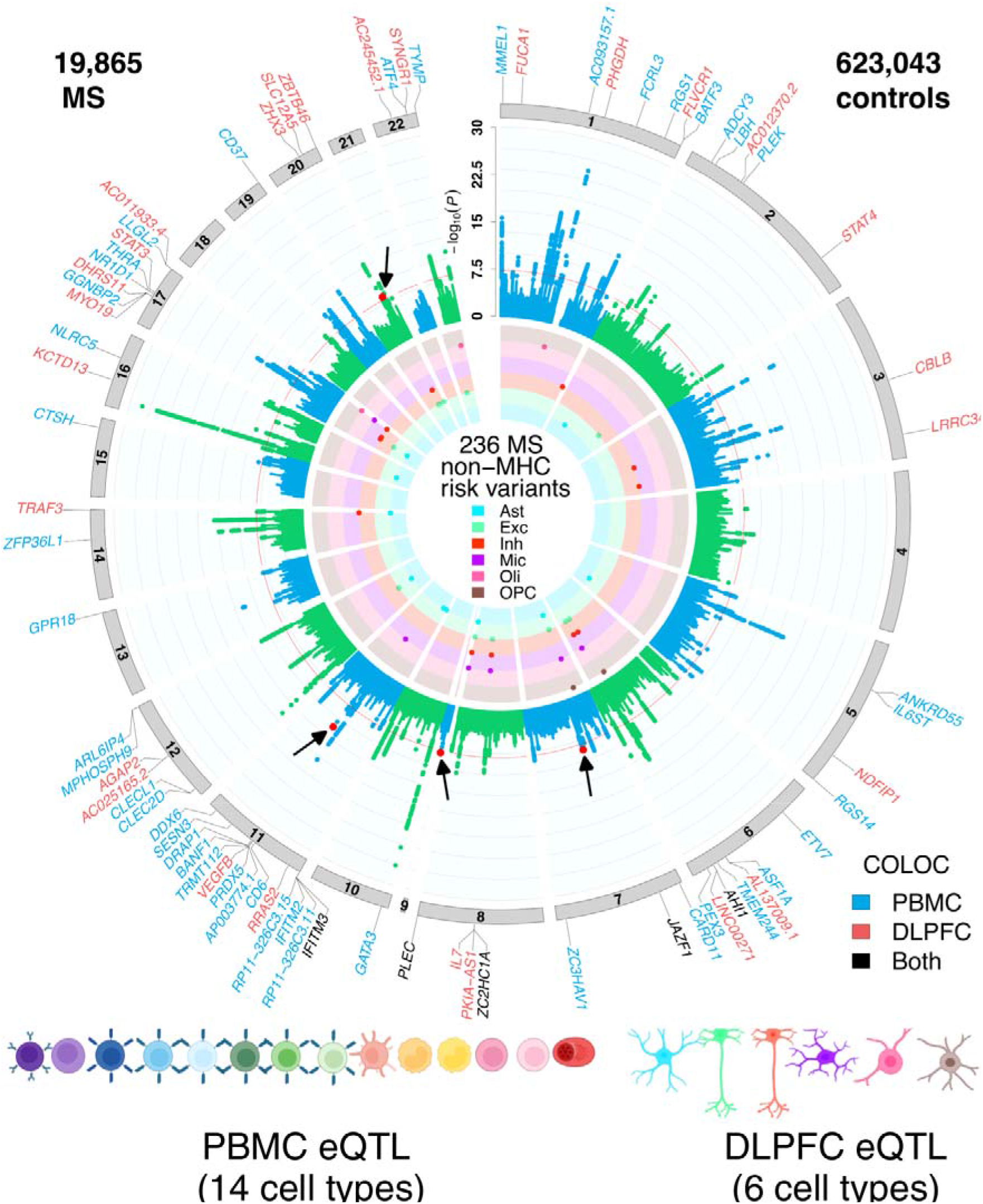
Circular presentation of loci associated with multiple sclerosis identified in European ancestry. The −log10(P) for genetic association with multiple sclerosis are arranged by chromosomal position, indicated by alternating blue and green points. Association P-values are truncated at P[<[1[×[10^−30^. Genome-wide significance (P[<[5[×[10^−8^) is indicated by the red line. Genes showing coloc effects with DLPFC cell types are highlighted in red, and the genes showed coloc effects in PBMC cell types are highlighted in blue, and the shared coloc genes annotated with black. The inner circle indicates MS-loci that co-localize with DLPFC QTL, colored by cell type. Color keys representing cell types are indicated in the plot center. Chromosomes are indicated by numbered panels 1–22.

**Table 1.**
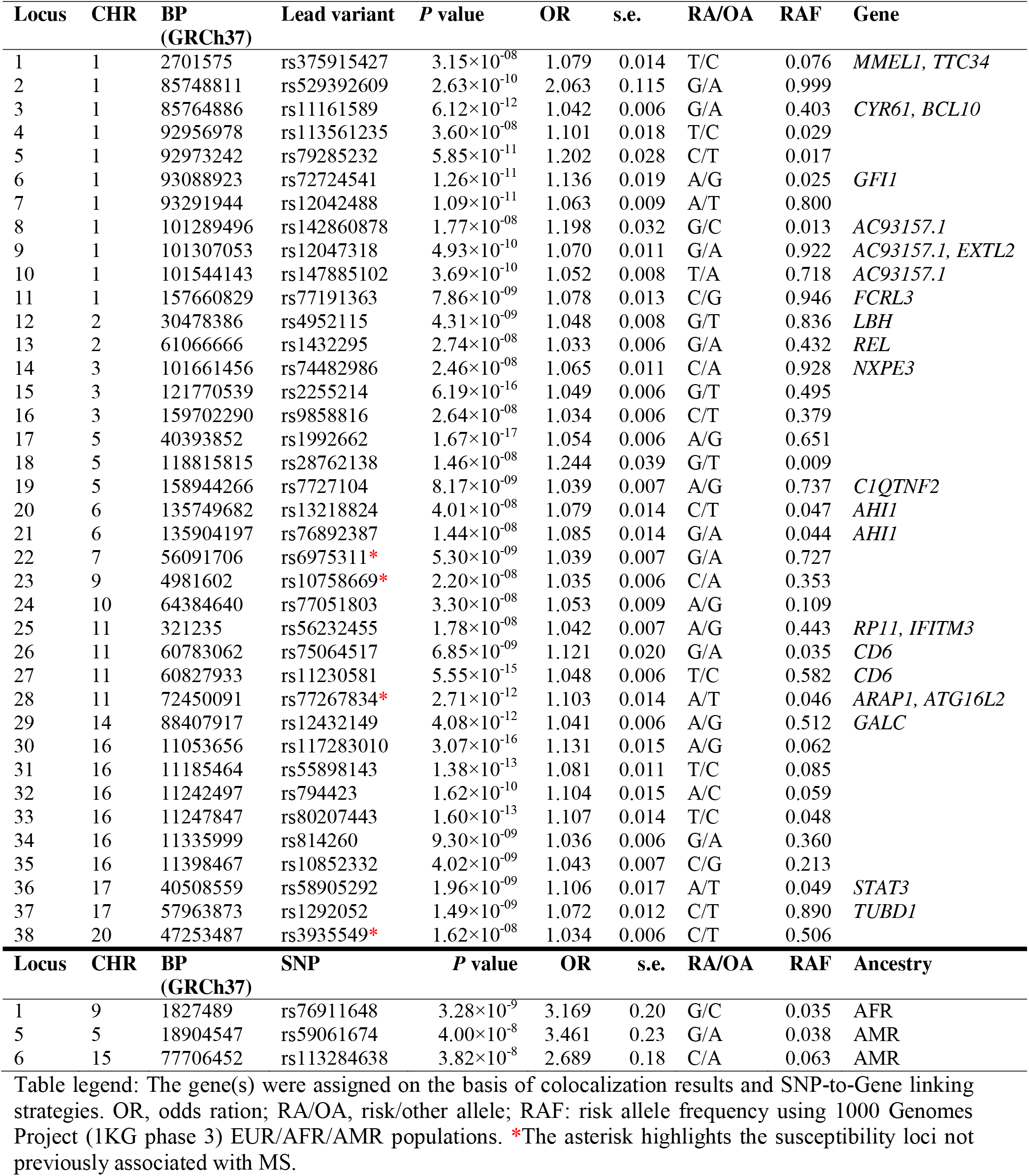
GWAS meta-analysis uncovers 38 additional MS susceptibility variants in EUR, one in AFR and two in AMR.

Previous studies reported that multiple MS loci harbored more than one statistically independent effect that met a genome-wide significance threshold (*1*). This pattern was also observed in our updated list of MS risk variants, where multiple independent associations were found at many loci, such as the *DDX6-CXCR5* locus (**Supplementary fig. S1**), which has also been implicated in other autoimmune diseases. For example, the variant rs12365699-G^,^ located in the *DDX6- CXCR5* locus, increased the risk of rheumatoid arthritis and lupus (*25–27*).

### Multi-ancestry GWAS meta-analysis for MS

Although the total number of AFR (614 cases and 62,044 controls) and AMR (352 cases and 44,133 controls) ancestry individuals identified by us across the three biobanks was large for an MS study, it remains modest for a GWAS. Our prior study of European ancestry participants required 1,000 MS cases to yield two loci meeting a threshold of genome-wide significance (*28*). Nonetheless, we completed separate GWAS for these two populations aiming to identify ancestry-specific loci. Surprisingly, one new locus is genome-wide significant among AFR participants (rs76911648), its minor allele frequency (MAF) in EUR (MAF=0.013) is lower compared to AFR (MAF=0.035), and two new SNPs are significant among AMR participants (rs59061674, rs113284638) (**Table 1**, **fig. S2**), where rs59061674 has a lower MAF in EUR (MAF=0.010) than AMR (MAF=0.038), while the rs113284638 showed a slightly higher MAF in EUR (MAF=0.072) than AMR (MAF=0.063) . Given small sample size, these results should be viewed cautiously; in participants of European ancestry, none of these three SNPs have a p<0.05. Further, they are not in LD with one of the significant SNPs described above. There are no additional non-European ancestry datasets available for replication, so these results will require validation in future more diverse cohorts.

To be thorough, we next performed a multi-ancestry meta-analysis of a total of 20,831 MS cases (>40% increase in the number of MS cases used in the previous GWAS (*1*)), and 729,220 control participants using two methods: a multi-ancestry meta-regression implemented in MR-MEGA (*29*) and a random effects model implemented in PLINK v1.9 (*30*). No additional loci became significant in this slightly larger meta-analysis. When we took the list of 236 significant SNPs from the EUR meta-analysis, 184 SNPs were available in AFR, and 18 of these SNPs showed some evidence for replication among AFR participants (nominal P<0.05, 14 with the same effect directions). In addition, 189 of the 236 SNPs could be tested in AMR participants, and 11 SNPs showed some evidence of association (P<0.05, 9 with the same effect directions) (**Supplementary table S3**). Thus, while dedicated studies of non-European populations are sorely needed, our results suggest that certain findings from European-ancestry meta-GWAS are also relevant to AFR and AMR populations, consistent with earlier studies (*31, 32*).

### Susceptibility alleles overlap between MS and other autoimmune diseases

Next, we assessed the extent to which MS susceptibility was shared with other diseases. We assembled a list of SNPs that reached genome-wide significance (p<5×10^-8^) in at least one of 12 autoimmune diseases using their publicly available genome-wide summary statistics (see details in Methods) (*33–40*). Adding these SNPs to those meeting a threshold of genome-wide significance in our MS analysis, 32,901 SNPs were retained for a cross-disease analysis (**Fig. 3A**). A single risk allele (rs3184504-T), a nonsynonymous SNP in the *SH2B3* gene, exhibited the highest level of pleiotropy with concordant risk associations shared across seven autoimmune diseases (**Supplementary table S4**), including multiple sclerosis, psoriasis, lupus, type 1 diabetes, celiac diseases, inflammatory bowel disease and thyroiditis. In addition, we identified 7,849 SNPs with associations shared between at least two autoimmune diseases, and we see decreasing numbers of SNPs that have some evidence of association in 3 more diseases, including 5 SNPs that may have a role in 6 diseases.

**Fig. 3.**
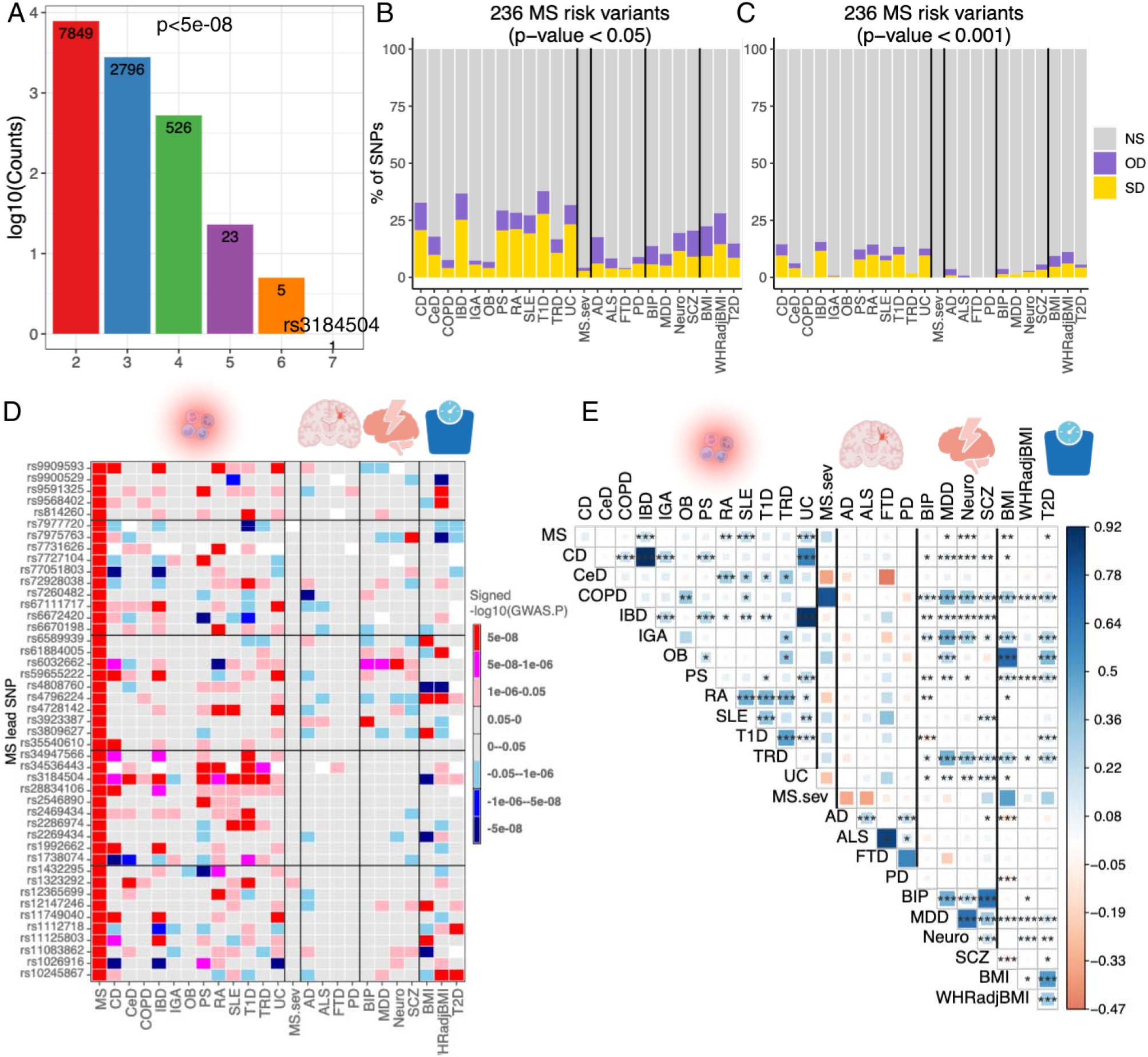
Overlap of the genetic architecture of multiple sclerosis 1032 with other diseases/traits. **(A)** The number of SNPs that reached genome-wide significant (P[<[5[×[10^−8^) and were shared across 12 autoimmune diseases. **(B, C)** Percentage of non-major histocompatibility complex SNPs of MS severity, 12 inflammatory/4 neurodegenerative/4 psychiatric/3 BMI-associated diseases/disorders/traits that are not statistically significant (NS), or significant in the same direction (SD) or the opposite direction (OD) in the current 236 MS risk variants using two P-values cut-off (p<0.05 and 0.001). Cell types are ordered alphabetically from left to right. **(D)** The comparison of 45 MS risk variants with other 24 diseases/traits, the colors represent effect directions and p values. White color denotes SNPs that were not detected in the corresponding phenotypes. **(E)** Genetic correlation estimated across MS and other 24 diseases/traits. The areas of the squares represent the absolute value of corresponding genetic correlations. After FDR correction for 325 tests at a 5% significance level, genetic correlation estimates that are significantly different from 0 are marked with an asterisk (*.01 < pFDR < .05; **.001 < pFDR < .01; ***pFDR < .001). The blue color denotes a positive genetic correlation, and the red color represents a negative genetic correlation.

To extend this analysis and better understand the pleiotropy of our MS variants, we next evaluated the behavior of our updated list of 236 MS susceptibility SNPs (**Supplementary table S2**) in the results of other GWAS, including MS severity (*2*), 12 autoimmune diseases (*33–40*), four neurodegenerative diseases (*41–44*), four psychiatric disorders/traits (*45–48*), and three metabolic traits (*49, 50*) (**Supplementary table S5**). The analysis was run twice, using either a nominal significance level (p<0.05) or a slightly more conservative threshold of p<0.001. We partitioned the results into three groups: (1) SNPs showing the same direction of effect as MS in the other disease/trait, (2) SNPs showing the opposite direction with these phenotypes, and (3) SNPs that did not meet the threshold of significance (**Fig. 3B & C**). T1D and IBD had the most potential associations for our MS SNPs, with 87 and 73 of SNPs meeting a nominal threshold of significance (**Fig. 3B**). In both cases, there was a clear skew for the sharing to occur in the same direction of effect, but a quarter of the MS variants had a flipped direction of effect in the other diseases. This pattern held true for the other autoimmune diseases and for the higher threshold of significance (**Fig. 3C**), consistent with patterns seen in earlier studies (*18*). The extent of sharing is dependent, in part, on the size of the GWAS for the non-MS trait, as some diseases still have relatively small GWAS or are underpowered, such as the MS severity GWAS, which returned only one significant locus (*2*).

Interestingly, we see a fair amount of sharing with the neuropsychiatric traits, more than with the neurodegenerative diseases. Alzheimer’s disease is intriguing, given an apparent excess of inverse effects in the shared SNPs with MS (p<0.05). The metabolic traits also harbor a notable amount of sharing. However, the direction of these variants seems relatively random, with ∼50% of the variants having an inverse effect relative to the MS risk. Under the more stringent statistical significance cutoffs (p<0.001), few of the MS SNPs were associated with the other traits, but the pattern among the autoimmune diseases was the same (**Fig. 3C**).

**Fig. 3D** presents the same results in a more granular form, where we filtered the MS risk variants that showed genome-wide significance in at least one of the 24 phenotypes utilized here: for example, the rs3184504-T allele in the *SH2B3* locus consistently shows increased risk in MS, lupus, celiac disease, thyroiditis, psoriasis, RA, and IBD (**Supplementary table S4**). We gain an appreciation of the complexity of the mechanisms of MS susceptibility: while a good portion of the variants clearly affect some aspect that yields a propensity to develop an autoimmune response, the substantial number of inverse effects highlight that the role of certain immune pathways is disease-specific. One example of this complexity is the *STAT3* locus, in which rs1026916 reaches p < 10^−28^ in MS (**Fig. 3D**) and has substantial evidence of being involved in psoriasis in the same direction, but this variant has attained genome-wide significance in IBD, UC, and CD in the opposite direction of effect relative to MS. Despite many shared autoimmune SNPs with MS, 50/236 were specific to MS at the most comprehensive threshold (p<0.05) across the 12 autoimmune diseases, and 27 were specific to MS among all the phenotypes we tested.

With the genome-wide summary statistics collected above, we then obtained the genetic correlations estimate from MS for the 325 pairwise combinations among the 25 phenotypes and compared the results to the LD score regression (LDSC) estimates (**Fig. 3E**, **Supplementary table S6**) using an imputed reference panel including 1,217,312 quality-controlled HapMap3 SNPs (*51*). This analysis suggests that MS is most similar to UC (rg=0.250, p-value=5.47×10^-09^), SLE (rg=0.221, p-value=1.00×10^-04^, IBD (rg=0.194, p-value=3.60×10^-06^) and RA (rg=0.150, p-value=1.50×10^-03^), which is consistent with earlier studies (*52, 53*). Interestingly, we also found significant positive correlations between MS and neuroticism (rg=0.090, p-value=2.00×10^-04^). MS severity did not show any significant correlations with the traits we tested. This MS severity study is probably underpowered, and we will need larger studies to truly explore the possibility of shared genetic architecture between MS severity and other inflammatory and neurodegenerative diseases. Notably, IgA nephropathy, COPD, thyroiditis, IBD, and CD had a significant genetic correlation with the psychiatric disorders/traits we tested (**Fig. 3E**).

### Polygenic score for multiple sclerosis

Polygenic risk scores have emerged as tools with which to capture an individual’s inherited disease susceptibility. They may be useful for stratifying individuals in clinical trials, and for guiding primary prevention and management of individuals at high genetic risk for MS (*54*). A genome-wide polygenic score (GPS) may also be used for discovery of the shared genetic architecture between MS and other unsuspected traits beyond inflammatory diseases.

We rigorously approached the construction of such a score using our non-MHC SNPs. We developed our initial model in the combined IMSGC, UKBB, and AoU datasets. We reserved the eMERGE-III dataset to test the model. The flowchart summary of our analytical approach is provided in **Figure 4A** (see details in Methods). The GPS for MS was tested with adjustment for age, sex, genotyping batch, and genetic ancestry. As shown in **Table 2**, the GPS was strongly associated with the risk of MS in the independent testing cohort of European ancestry, with an overall odds ratio (OR) per standard deviation of the GPS of 1.70 (95%CI:1.52-1.91, P=1.37×10^D19^). The participants in the top 1% vs. the remaining 99% of MS-GPS had more than a 7-fold increased MS risk (95% CI: 4.37-12.00, *P* = 1.60×10 ¹ ). We additionally validated the risk score in two smaller testing cohorts of AMR and AFR ancestry. Although the magnitude of effect was decreased, the GPS was significantly associated with MS in both cohorts. The OR per standard deviation of the GPS was estimated at 1.46 (95%CI 1.10-1.94, P=8.57×10 ^03^) for AMR ancestry and 1.26 (95%CI 1.07-1.49, P=5.64×10 ^03^) for AFR ancestry (**Table 2**). Thus, while dedicated efforts in these populations are sorely needed to further improve the GPS performance, the current GPS is already validating across major ancestral populations found in North America.

**Fig. 4.**
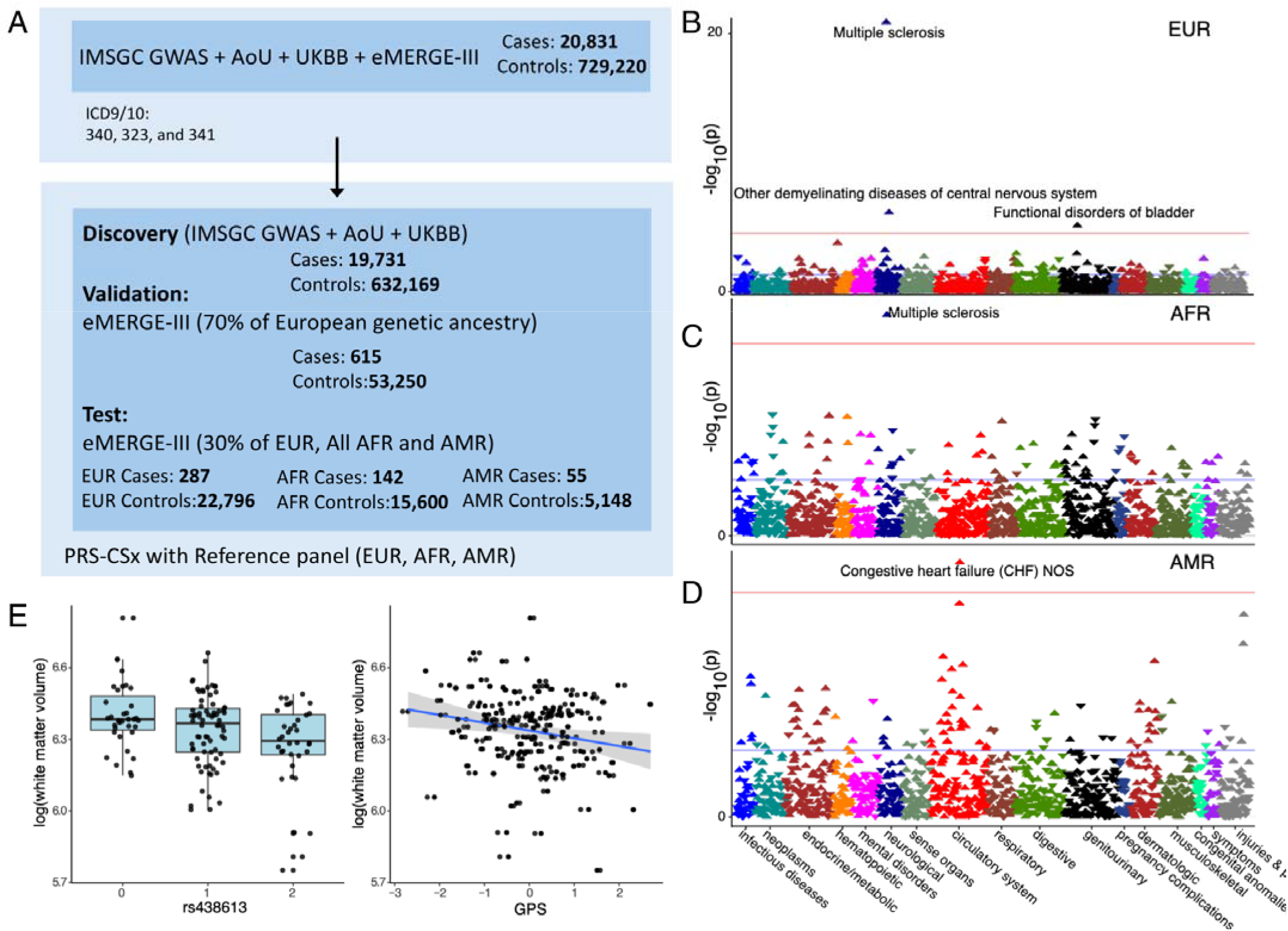
Workflow for the analaysi of MS GPS. **(A)** The MS GPS was developed using GWAS summary statistics from the IMSGC, All of Us (AoU), and UK Biobank (UKBB). Optimization was performed using 70% of European ancestry participants from eMERGE-III. GPS performance was validated in the remaining 30% of eMERGE-III participants of EUR and all AMR and AFR. **(B, C, D)** PheWAS results are shown for European (N = 23,121), African-American (N = 15,863), and Latino (N = 5,224) participants. The analysis includes combined data from eMERGE participants with both genotype and phenotype information. Logistic regression was used, adjusting for age, sex, batch, and ancestry. Effect estimates and two-sided P-values were reported. Red horizontal lines indicate the phenome-wide significance threshold, adjusted for multiple testing (P = 2.8 × 10[[). The Y-axis represents -log10(P-value), and the X-axis displays system-based phecode groupings. Upward-pointing triangles indicate increased odds for a given phecode, while downward-pointing triangles indicate reduced risk. **(E)** Boxplot diagram depicts the genetic effect of rs438613 with a significant association with white matter volume. The scatter plot displays the pattern of MS GPS in relation to white matter volumes.

**Table 2.** Performance metrics for the genome-wide polygenic score (GPS) in MS.

| eMEGRE-III<br>(Ancestries) | Case/control | OR per SD (95% CI),<br>P-value | AUC (Crude) | PRS Threshold | Odds ratio (95% CI), P value |
| --- | --- | --- | --- | --- | --- |
| **EUR (30%)**<br>**Genetic Ancestry** | 287/22,796 | 1.70 (1.52-1.91),<br>$P=1.37 \times 10^{-19}$ | 0.7217 (0.6398) | Top 20% vs. other 80% | 2.62 (2.06-3.34), $P=3.71 \times 10^{-15}$ |
| | | | | Top 10% vs. other 90% | 3.00 (2.28-3.95), $P=3.11 \times 10^{-15}$ |
| | | | | Top 5% vs. other 95% | 4.14 (3.03-5.65), $P=4.45 \times 10^{-19}$ |
| | | | | Top 2% vs. other 98% | 5.05 (3.3-7.75), $P=9.45 \times 10^{-14}$ |
|  |  |  |  | **Top 1% vs. other 99%** | **7.24 (4.37-12), $P=1.60 \times 10^{-14}$** |
| **AFR Genetic**<br>**Ancestry** | 142/15,600 | 1.26 (1.07-1.49),<br>$P=0.00564$ | 0.7458 (0.5543) | Top 20% vs. other 80% | 1.45 (0.99-2.13), $P=0.057$ |
| | | | | Top 10% vs. other 90% | 1.53 (0.95-2.46), $P=0.079$ |
| | | | | Top 5% vs. other 95% | 1.70 (0.93-3.11), $P=0.087$ |
| | | | | Top 2% vs. other 98% | 1.73 (0.69-4.32), $P=0.241$ |
|  |  |  |  | **Top 1% vs. other 99%** | **2.70 (0.96-7.61), $P=0.060$** |
| **AMR Genetic**<br>**Ancestry** | 55/5,148 | 1.46 (1.10-1.94),<br>$P=0.00857$ | 0.7524 (0.5526) | Top 20% vs. other 80% | 1.41 (0.73-2.73), $P=0.308$ |
| | | | | Top 10% vs. other 90% | 1.52 (0.63-3.66), $P=0.346$ |
| | | | | Top 5% vs. other 95% | 2.14 (0.74-6.14), $P=0.159$ |
| | | | | Top 2% vs. other 98% | 5.99 (2.02-17.8), $P=0.001$ |
|  |  |  |  | **Top 1% vs. other 99%** | **9.87 (2.75-35.5), $P=4.44 \times 10^{-4}$** |

We then assessed phenome-wide associations of this GPS in a PheWAS based on eMERGE participants who were not included in the GPS design. This approach offers a complementary strategy to assess for unsuspected shared genetic architecture with a range of clinical traits across the entire phenome. In the well-powered analysis of participants with European ancestry, the GPS association with MS was strongly replicated (OR=1.97, 95%CI:1.71-2.27, P=1.35×10^-21^, **Fig. 4B**). We also found a GPS association with “other inflammatory demyelinating diseases” (OR=1.67, 95%CI:1.36-2.04, P=7.33×10^-07^, **Fig. 4B**). This poorly defined diagnostic group may harbor certain individuals with MS and contains conditions that share symptomatology with MS but have different immune mechanisms. Thus, there may be some overlap in genetic architecture with these less common entities. The association with “functional disorders of the bladder” (OR=1.22, 95%CI:1.12-1.34, P=7.59×10^-06^, **Fig. 4B**) was likely related to the fact that bladder dysfunction represents a common symptom of MS. No other diagnostic category was significantly associated with the MS GPS, suggesting that our GPS is fairly specific to MS.

In the smaller AFR dataset, the association of the GPS with MS was also phenome-wide significant (OR=1.62, 95%CI:1.31-2.01, P=7.55×10^-06^, **Fig. 4C**), consistent with the earlier dedicated analysis. In the AMR cohorts with fewer cases, the association with MS was only nominally significant (OR=1.51, 95%CI:1.01-2.26, P=0.04, **Fig.4D**) likely due to low power. The GPS was also associated with “Congestive heart failure (CHF) NOS” (OR=1.28, 95%CI:1.15-1.43, P=1.08×10^-05^, **Fig.4D**). MS has been reported to be linked to a higher risk of cardiovascular disease, including congestive heart failure (*55*), but given small sample size of the AMR cohort, and the absence of this association in AFR and EUR cohorts, this association may be spurious. We conclude that the GPS is associated with MS across different ancestral populations, but its predictive performance remains lower in non-European populations.

Finally, we have tested our GPS in MS patients with magnetic resonance imaging (MRI) measurements (gray matter, white matter, and cerebrospinal fluid), the Expanded Disability Status Scale (EDSS), and genotype information using the data selected from the Comprehensive Longitudinal Investigation of Multiple Sclerosis at the Brigham and Women’s Hospital (CLIMB) study (*56*) (see details in Methods). A linear regression model was used to examine the associations between the GPS and brain tissue compartments adjusted for age at visit, sex, and top three ancestry PCs. We observed a nominally significant association between the GPS and lower white matter volume in the MS patients (p-value = 0.03) (**Fig. 4E**). However, no significant associations were found between MS-GPS and other MRI measurements.

### Functional characterization of MS variants using cell-type specific brain and blood eQTL

Prior systematic evaluations of functional consequences of MS susceptibility variants (*1, 2*) had revealed that MS SNPs affected gene expression in peripheral immune cells and microglia (a myeloid cell type that integrates the neurectoderm early in development). While some targeted investigations looked at astrocytes in relation to molecular pathways present in many cell types, there has been few systematic evaluation of MS genetic effects specific to CNS cell types in relation to MS susceptibility or severity (*2, 57–61*). Thus, we accessed our recent atlas of CNS cell type-specific eQTL effects generated from well-powered set of frozen postmortem human brain samples collected from the same brain region, the dorsolateral prefrontal cortex (*62*). Further, we accessed a publicly available resource derived from PBMC samples (*63*) to map the effects of MS variants on peripheral immune cells as a contrast and to assess cell-type specificity of the functional consequences. Using these two references, we identified those MS susceptibility variants that are found in the vicinity of an eQTL in each of the tested blood and brain cell types and then assessed whether the two effects co-localize using Coloc (v5.1.0) (see **Methods**). The results are shown in **Figure 5A (Supplementary table S7)** where, as expected, there are several colocalized effects (PP.H4>0.8) among blood-derived cells; this is consistent with prior reports that naïve T cells harbor the most of these MS-related functional consequences (*64, 65*). Most of these effects are shared among several cell types, but some – such as *NR1D1* and *MMEL1* – appear specific to naïve T cells (amongst the cells surveyed here). Further, we now demonstrate colocalization with microglial eQTL, which had been suspected from prior analyses that uncovered enrichment of microglial genes amongst genetically implicated MS susceptibility genes (*1, 6*).

**Fig 5.**
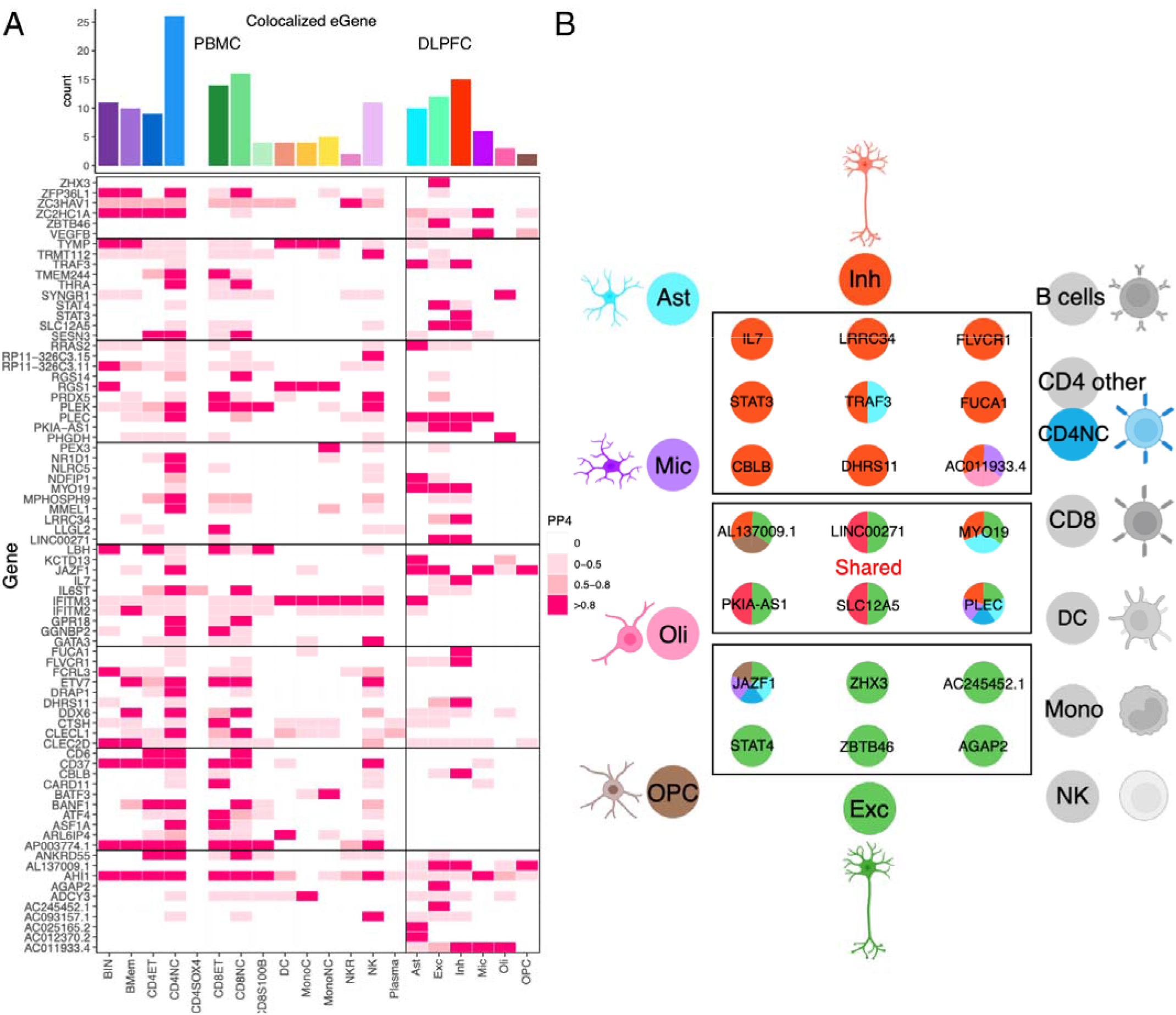
Overlap of the results from PBMC/DLPFC eQTL and GWAS of MS. **(A)** Heatmap reports the PP.H4 of the Coloc method, which assumes that GWAS and eQTLs share a single causal SNP. The rows report the overlap for individual gene and SNP pairs; the columns report the PP.H4 score in each of our cell types. The color of each square is based on the code found to the right; the darker color denotes higher confidence that the same variant influences susceptibility and gene expression in that cell type. The top bar chart shows the number of colocalized eGenes with high confidence (PP.H4[>[0.8) in each cell type. **(B)** Cartoon illustration summarizes the colocalization effects of neurons compared to the 18 cell types included in our analysis, colored by cell type.

However, the most interesting new set of results involves the cell types that derive from the neurectoderm: the glial and neuronal cells. In our reference, the most numerous cell types are excitatory neurons; they also have the largest transcriptome and, hence, have the most eQTL effects compared to other CNS cell types that are less frequent in the cortex (*62*). Despite this, inhibitory neurons harbor the most eQTLs that colocalize with MS susceptibility variants of any CNS cell type (n=15) (**Fig. 5A & B & S3**); this is more than the resident immune cells, the microglia (n=6). Further, seven of these functional consequences to MS variants are unique to inhibitory neurons. We also see five other variants that have functional consequences only in excitatory neurons. Thus, neuronal cells seem to play an important role in the earliest events leading to the onset of MS. **Figure 6A&B** zooms into two MS loci, *STAT3* and *IL7*, illustrating the co-localization of susceptibility and expression effects. These are well-studied cytokine-related genes involved in amplification of immune responses, with evidence that IL7-driven signaling occurs, in part, through *STAT3*. Our comparative assessment of blood and brain cells indicates that these two functional consequences of MS variants may be mechanistically related and unique to inhibitory neurons. They may provide a bridge between the peripheral leukocyte-driven propensity for autoimmunity and the targeting of the CNS by peripheral immune dysfunction, as neuronal cells respond differently to inflammatory stimuli. Further work is needed to understand how these two functional consequences intersect with the other neuronal-specific effects (in excitatory as well as inhibitory neurons).

**Fig. 6.**
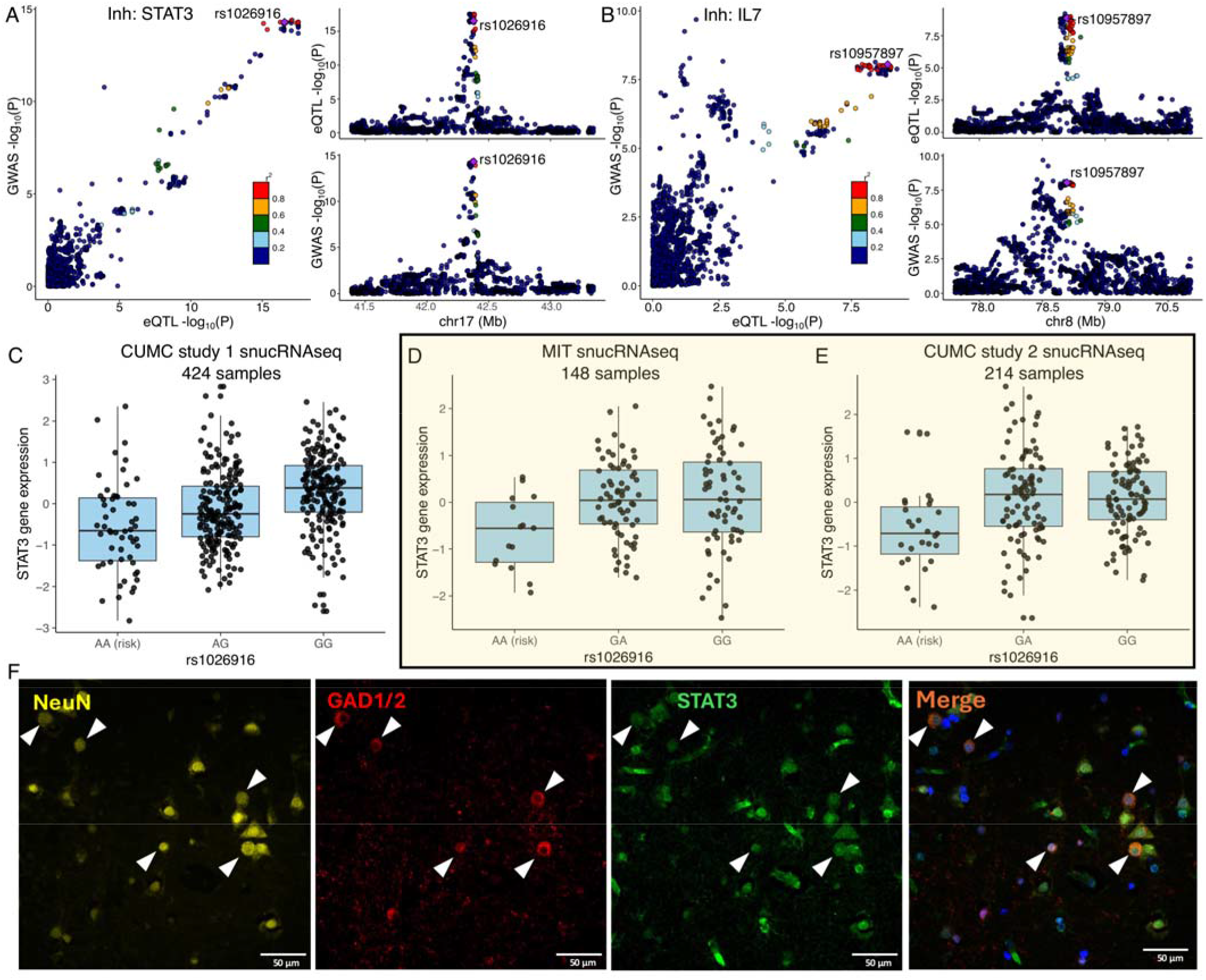
Examples of COLOC results. **A), B)** The locus-compare scatter plot for the association signals at *STAT3* and *IL7* in the inhibitory neurons. **(C, D, E)** Expression quantitative trait loci (eQTL) box plots of associations between genotype rs1026916 and *STAT3* expression in inhibitory neurons using snucRNAseq data from Fujita et al. (CUMC study 1), Mathys et al. (MIT cohort), and our in-house multiome datasets (CUMC study 2). **(F)** Immunohistochemistry of DLPFC in human MS brain tissue, stained for STAT3 (green), GAD1/2 (red), and NeuN (yellow), with DAPI (blue) to visualize nuclei. Expression of STAT3 was observed in NeuN+GAD1/2+ neurons. White triangles highlight the colocalization of DAPI, STAT3, GAD1/2, and NeuN. Scale bar, = 50 μm.

While neurons harbor the most functional consequences of MS variants, each of the glial cell types harbor some such effects, including some that are specific to astrocytes (*KCTD13* and *RRAS2*) and oligodendrocytes (*PHGDH* and *SYNGR1*), for example. A previous report implicated an MS variant near the *NFKB1* gene in altered immune responses in astrocytes; however, this SNP is not found to alter gene expression in our brain datasets (*66*). We note that the pathognomonic feature of multiple sclerosis at its onset is the presence of inflammatory demyelination, which targets the myelin sheath produced by oligodendrocytes. Thus, while some of the MS loci may finally connect the peripheral immune dysfunction to a well-validated target cell type, many more loci implicate neuronal cells, and this may provide insights into the neurodegenerative component of the disease, which is apparent as brain atrophy early on (*67, 68*) but presents clinically only much later.

### Replication of colocalized eQTL & epigenomic assessment

We accessed additional single-nucleus datasets, and, as shown in **Figure 6C-E**, and the *STAT3* RNA expression effect in inhibitory neurons is robust, being reproducibly found in two other datasets (*69*). The rs1026916^A^ risk allele is associated with decreased gene expression using our original dataset (CUMC study 1) (*62*), data from colleagues at the Massachusetts Institute of Technology (MIT) (*69*), and a new snucRNAseq dataset (CUMC study 2). In addition, we identified eQTL-eGene effects in multiple corresponding cell types (**Supplementary table S8**). For example, rs4896153 is an eQTL associated with *AHI1* in microglia, and rs6032662 is associated with *SLC12A5* in both excitatory and inhibitory neurons.

Reviewing reference epigenomic profiles (*70*), we found that most of our top prioritized variants (**Figure 5 and Supplementary Table S7**) are not located in segments of open chromatin in the cell types implicated by the eQTL analyses. However, one SNP, rs3923387, tags a genetic effect near the *PLEC* gene that influences (1) MS susceptibility, (2) the accessibility of chromatin in a nearby chromosomal segment (GRCh37 chr8:145034681-145035181) in microglia (colocalization of MS susceptibility PP.H4=0.84), and (3) the expression of the *PLEC* gene in the same cell type (as shown in Figure 5, colocalization of the ATAC QTL and eQTL PP.H4=0.93 (**Supplementary fig. S4**). This result illustrates the next phase of our consortium’s work, generation of improved, cell-resolved, multi-omic data to map the propagation of effects from the MS susceptibility variants.

Finally, to extend the narrative, we also confirmed the expression of STAT3 protein in inhibitory neurons using immunofluorescence in DLPFC tissue sections of a post-mortem MS individual obtained from the New York Brain Bank (NYBB). We observed that STAT3 is expressed in GAD1^+^GAD2^+^ inhibitory neuron cells, with 2.8% of these neurons showing elevated STAT3 expression (>2SD) (**Fig. 6F and S5**). However, no significant differences in neuronal morphology, including compactness and shape, were observed between STAT3-expressing inhibitory neurons and those lacking STAT3 expression.

## Discussion

In an updated MS GWAS analysis of 19,865 MS cases with genome-wide genotype data, we identified 38 novel MS risk variants and four novel genomic loci involved in MS susceptibility. Combined with SNPs generated from previous studies (*1*), our consortium has reported a total of 236 independent non-MHC MS risk variants identified in participants of European ancestry. We have also conducted a multi-ancestry MS GWAS, including AFR and AMR ancestry participants. Although the sample size of the diverse participants is small, we uncovered one locus that reached genome-wide significance among AFR participants and two loci among AMR participants. These results should be considered cautiously until further evidence of replication emerges, given the small size of their discovery analyses. Our rigorously derived GPS provides a new tool for the community to investigate the role of genetic predisposition to MS in other datasets and contexts. Interestingly, while it requires additional optimizations for use in non-European populations, our results suggest that the current version already has some predictive capacity among individuals of AFR and AMR ancestry, consistent with earlier reports (*71, 72*). There is a pressing need for larger studies in non-European ancestry groups to ensure that any future clinical utility is broadly applicable. Given the strong but complex role of the MHC in MS, inclusion of susceptibility variants from that region will further improve the prediction in European populations but may be less informative in diverse population given the rapid and copmplex evoluation of the MHC which harbors many population-specific effects.

Using our updated MS results, we sought to classify our susceptibility variants functionally. In one approach, we accessed the results of other GWAS to identify those variants that may affect susceptibility by altering more general mechanisms that lead to a propensity for autoimmunity. This hypothesis is consistent with epidemiological studies reporting a higher prevalence of other autoimmune diseases in persons with MS, such as T1D, thyroid disease, and inflammatory bowel disease (*73, 74*) as well as the existence of families with members affected by different autoimmune diseases (*75*). However, the story is not that simple, as there does not appear to be a clear “global genetic risk for autoimmunity”: The rs3184504 variant in the *SH2B3* locus offers a good illustration, as its risk allele “T” was found associated with increased risk of celiac disease, IBD, MS, psoriasis, lupus, T1D and thyroiditis. The *SH2B3* gene encodes the Src homology 2 adaptor protein 3, which regulates inflammation, immunity, and blood cell production. Certain genetic variants of *SH2B3* can cause it to fail to control an overactive immune response, which can lead to autoimmunity (*76*). It was also reported to be associated with immunoglobulin levels and multiple other non-immune traits; it displayed a high degree of pleiotropy, being associated with 79 different GWAS traits (*77*). Overall, our results support shared autoimmune mechanisms (*52*), where we show that a substantial proportion of shared loci harbor pleiotropic effects influencing risk to MS and other autoimmune diseases.

We thus found that 186 of the 236 variants have some evidence of association with another autoimmune disease using the most inclusive threshold. This suggests that the remaining 50 variants may have a role in other processes that relate to targeting the propensity for an autoimmune process towards the target organ, in our case, the brain and spinal cord. Prior work had clearly demonstrated that the peripheral immune system harbors the functional consequences of many variants. While CD4^+^ T cells were strongly implicated, all other bone marrow-derived cells and microglia were also found to harbor at least some of the effects of susceptibility variants (*1, 2*) in these analyses. The role of CNS cells was unclear, with a potential but ambiguous association with *SLC12A5* expression in brain transcriptomic data and functional consequences of the six MS variants in astrocytes that perturbed the NF-κB pathway. This pathway is also implicated in many immune cells, and current MS treatments are found to be directly or indirectly linked to NF-κB pathways, modulating both the innate and adaptive immune system in patients (*78–80*).

Here, our co-localization analysis showed that CD4^+^ Naïve T cells harbor the largest number of cases where the same variant influences MS susceptibility and RNA expression, consistent with previous studies. Surprisingly, we found that inhibitory neurons showed the most colocalization signals among CNS cell types, followed by excitatory neurons, astrocytes, and microglia, and most of the colocalized signals in neurons are unique to this cell type (when compared to cortical and bone-marrow-derived cells). For example, *STAT3* and *IL7* illustrate loci with evidence of co- localization of susceptibility and expression effects only in inhibitory neurons. These are well-studied cytokine-related genes that are involved in the amplification of immune responses, with evidence that *IL7*-driven signaling occurs, in part, through *STAT3* (*81*). Thus, these two loci implicate a specific molecular pathway in the onset of MS through perturbation of neuronal function. Another example is *ZHX3* in excitatory neurons, *ZHX*3 is a member of a family of transcriptional repressors that are involved in neural progenitor maintenance, hematopoietic cell development, and differentiation. Dysfunction of *ZHX* family members is linked to the development and progression of neurological disease (*82*). Our comparative assessment of blood and brain cells, therefore, prioritizes a subset of MS variants that implicate CNS parenchymal cells in disease onset. Clearly, perturbed pathways that lead to a propensity to autoimmune reactions are interacting with perturbed immune responses in neurons and glial cells to initiate autoreactive cells that lead to both recurrent bouts of inflammatory demyelination and a slowly progressive neurodegenerative process that remains poorly understood. The predilection of inhibitory neurons as a target for these risk variants is intriguing, particularly given the recent report that inhibitory neurons appear to be lost preferentially in the MS brain (*83*). With our observations, we can now generate hypotheses to explore the downstream molecular and functional changes elicited by the variants in the cell type in which they are implicated. The role of the adaptive immune system is well established in MS, while CD8^+^ T cells are most abundant in the white matter of MS brain (*84*). The CD4^+^ T cells probably play a role that is as important given the convergence of genetic susceptibility effects in this cell type and earlier studies (*85*). Our data here suggest direct interactions between T cells and neurons or glia may be important to trigger the onset of MS through both classes of lymphocytes, elaborating a rich literature of immune responses expressed by neuroglial cells (*86, 87*). More broadly, it is likely that tissue-specific cells are likely to play a similar role in other inflammatory diseases.

In summary, these results advance our understanding of the biological etiology of MS, refocusing our efforts on understanding the onset of the disease to include specific molecular pathways in the brain. While most loci have functional consequences in a variety of immune cell types, our study prioritizes understanding the unsuspected neuronal contribution to the onset of MS. They alter our conceptualization and approach to primary prevention and treatment of MS, which may have to include interventions targeting the central nervous system pathways.

## Methods and Materials

### Study design

This cross-sectional study involves a combined analysis of the UKBB, eMERGE-III, and AoU cohorts. All participants provided informed consent to participate in genetic studies. Each cohort was first analyzed separately, and cohort-specific results were combined using fixed-effects meta-analysis.

### UK Biobank (UKBB)

The UKBB is a longitudinal cohort of individuals ages 40–69 years at enrollment, recruited between 2006 and 2010 across the United Kingdom (*19*). The individuals recruited to UKBB signed an electronic consent to allow the broad sharing of their anonymized data for health-related research. UKBB generated and released SNP microarray, exome sequence, and structured EHR data for 488,377 participants. The cohort is 54% female, with a mean age of 57 years, and the composition is 94% Europeans, 2% West or Southeast Asians, and 2% African ancestry by self-report (*19*) (**Supplementary table S1**).

### SNP microarray data

The details of the UKBB microarray genotyping, imputation, and quality control are available elsewhere (*19*). Briefly, using the UKBB Axiom Array (N = 438,427) and UK BiLEVE Axiom Array (N = 49,950), a total of 488,377 participants have been genotyped for 805,426 overlapping markers. The 1000 Genomes, UK10K, and Haplotype Reference Consortium (HRC) reference panels were used to perform genome-wide imputation using IMPUTE2 software (*88, 89*). We performed post-imputation quality control analyses as described in our previous work based on this dataset (*90*) retaining 9,233,643 common (i.e., Minor Allele Frequency (MAF) > 0.01), high-quality (imputation R2 > 0.80) variants for the purpose of GPS calculation. To eliminate any potential confounding by close familial relationships, we excluded cryptically related individuals (kinship coefficient > 0.0442) (*91*) from downstream analyses.

### Genetic ancestry analysis

We used the UKBB genotype array data for principal component analysis (PCA). We first pruned the genotype data using the plink command ‘--indep-pairwise 500 50 0.05’. We then used FlashPCA (*92*) based on 35,091 pruned variants. We merged the UKBB samples with 2504 participants of the 1000 Genomes Project (1KG phase 3) (*93*) and kept only shared variants between the two datasets. Then, we used a random forest machine learning based on 10 principal components to train ancestry classifiers using 1KG labeled data. Finally, we used the trained model to predict the genetic ancestry of the UKBB samples (**Supplementary fig. S6a**).

### All of Us (AoU)

The AoU research program launched recruitment in 2018 across 340 sites across the United States, and over 372,380 participants were enrolled by 2022. AoU combines participant-derived data from surveys such as self-reported health information, physical measurements, electronic health records, and biospecimens. We analyzed the AoU data on Workbench, a cloud-based environment (*21*). The second release data included N = 312,944 participants with complete SNP microarray, genome sequencing data, and phenotype information. The participants included 60% female, the mean age was 55 years, and consisted of 53% European, 4% Asian, and 21% Black/African American race by self-report. In addition, 17% of the cohort self-reported Hispanic/Latinx ancestry (**Supplementary table S1**).

### SNP microarray genotype data

All participants were genotyped with the Illumina Global Diversity Array (GDA). This microarray contains 1,904,679 SNVs and 44,172 indels. First, we performed genome-wide imputation analysis on the Workbench platform. Before imputation, we excluded all variants with MAF less than or equal to 0.005 (671,685 variants) or genotype missingness rate greater than or equal to 0.05 (41,526 variants). The genomic positions were lifted over from human GRCh38 to hg19 for 96% of SNPs. We then adopted the TopMed pre-imputation quality control (QC) pipeline to correct allele designations and remove poorly mapping variants (*94*). After QC, we used 1,191,468 variants for imputation. To reduce RAM usage and increase speed, we split the 312,944 subjects with microarray data into 8 equal batches and then imputed each batch separately. After pre-phasing with EAGLE v.2 (*95*), we imputed missing genotypes using the Minimac4 (*88*) and 1KG phase 3v5 (*93*) reference panel. A total of 43,371,225 autosomal variants were imputed in 312,944 individuals. We then merged the eight batches based on position using VCFtools software with the command ‘vcftools --gzvcf --positions --recode -- recode-INFO-all –stdout’. MAFs for the imputed markers were closely correlated (correlation coefficient (r) = 0.96) with the MAFs for the 1KG dataset.

### Genetic ancestry analysis

Similar to the UKBB data, we first pruned the genetic data using the command ‘--indep-pairwise 500 50 0.05’ in PLINK (*96*) and used N = 36,358 pruned variants for kinship and ancestry analysis. Using KING software (*91*), we removed 270 samples with pairwise kinship coefficients>0.35. We then merged our AoU samples with 1KG samples, kept only SNPs in common between the two datasets, calculated PCs for the 1KG samples, and projected each of our samples onto those PCs. We then used a random forest-based machine learning approach to assign a continental ancestry group to each AoU sample. Briefly, we trained and tested the random forest algorithm on 1KG subjects with known labels. We trained the random forest model using 10 PCs as a labeled feature matrix. Then, we used our trained random forest model to predict the genetic ancestries for the AoU dataset (**Supplementary table S1** and **Supplementary fig. S6b**).

### eMERGE-III

The eMERGE network provides access to electronic health record information linked to GWAS data for 102,138 individuals recruited in 3 phases (eMERGE-I, II, and III) across 12 participating medical centers from 2007 to 2019 (54% female, mean age 69 years, 76% European, 15% African-American, 6% Latinx and 1% East or southeast Asian by self-report) (*97, 98*). All individuals were genotyped genome-wide; details on genotyping and quality control analyses have been described previously (*97, 98*). All GWAS datasets were briefly imputed using the multiethnic Haplotype Reference Consortium panel on the Michigan Imputation Server (*99*). The imputation was performed in 81 batches. We included only markers with a MAF ≥ 0.01 and R2 ≥ 0.8 in ≥75% of batches post-imputation. A total of 7,529,684 variants were retained for the GPS analysis. For PCA, we used FlashPCA (*92*) on a set of 48,509 common (MAF ≥ 0.01) and independent variants (pruned in PLINK with the --indep-pairwise 500 50 0.05 command). The analyses were performed using a combination of VCFtools v.0.1.13 (*100*) and PLINK v.1.9 (*96*). Similar to UKBB and AoU, we defined the genetic ancestry for eMERGE based on random forest (**Supplementary fig. S6c)**.

### MS phenotyping and case-control definitions

The MS phenotype was defined using ICD codes from the UKBB, eMERGE-III, and AoU datasets. Cases were identified by at least one occurrence of the following ICD codes: ICD-9: 340, 323, or 341. Participants without any of these codes were classified as controls.

### Meta-GWAS

The MHC is the most gene-dense and most polymorphic part of the human genome. The region exhibits haplotype-specific linkage disequilibrium patterns, extreme structural variation and copy number variations, and an extremely high level of genetic diversity; the use of a single reference sequence to analyze GWAS data in this area is problematic (*101*). Therefore, the Extended MHC region (xMHC) is set aside in our meta-analysis (defined as the regions between *HIST1H2AA* and *RPL12P1* genes: chr6: 25,383,722-33,368,421Mb; ∼7.6Mb, GRCh37), resulting in ∼68,000 SNPs located in xMHC were removed for further analysis.

We first performed a meta-analysis using an inverse-variance-weighted fixed-effects model in METAL (version 2011-03-25) (*24*) combining UKBB, AoU, and eMERGE-III cohorts for European ancestry (5,063 MS cases and 596,340 controls), African-American (614 MS cases and 62,044 controls) and Hispanic American (352 MS cases and 44,133 controls) populations, respectively. In addition, another meta-analysis using METAL was performed exclusively for the European ancestry cohort, which included the GWAS summary statistics from the IMSGC discovery cohort (*1*), along with UKBB, AoU, and eMERGE-III (19,865 MS cases and 623,043 controls). A genome-wide significant locus was defined as the region around a SNP with P < 5 × 10^−8^, LD r^2^ > 0.1, within a 500-kb window, using the reference panel from phase 3 of the 1000 Genomes Project as the reference population.

Two models were used to conduct multi-ancestry meta-analyses (20,831 MS cases and 729,220 controls). Random effects models were performed using PLINK v1.9 (*96*), while a separate analysis was performed using MR-MEGA v0.2 (*29*). PLINK v1.9 was preferred over METAL due to its capacity to perform random effects analyses in parallel. A random effects model provides a more conservative framework that allows each study to have unique effects, as expected in different populations. MR-MEGA was also employed since it is well-powered to detect associations at loci with allelic heterogeneity. MR-MEGA models allelic effects as a function of axes of genetic variation that are derived from the input GWAS summary statistics. This method can result in reduced variant sets since it requires that variants have sufficient overlap between the input datasets (K > 3), where K is the number of inputs GWAS, in contrast to random effects models implemented in PLINK v1.9, which were limited to K > 2 to quantify heterogeneity accurately.

To identify novel genomic risk loci, LD blocks of independent significant SNPs (R^2^ > 0.1, ±500kb, 1KG phase 3) were merged into a single genomic locus if the distance between LD blocks was less than 250 kb. These loci were compared to the previous GWAS (*1*) to assess whether these regions were known to be associated with MS. There was no evidence of stratification artifacts or uncontrolled inflation of test statistics in the results from any cohort (λ GC = 1.02–1.14 **Supplementary fig. S2**).

### Conditional analysis

To identify secondary association signals, we used the program GCTA-COJO (*102*) to perform conditional analysis on the summary meta-analysis. GCTA-COJO (--cojo-cond) performs a secondary association analysis conditioned on discovered top variants; such conditional analysis is conducted with GWAS meta-analysis summary statistics rather than individual-level data of the full sample.

### Summary Statistics for Autoimmune Diseases and Other Traits

We downloaded complete summary statistics for autoimmune and inflammatory disease GWAS available in the NHGRI-EBI GWAS catalog( https://www.ebi.ac.uk/gwas/downloads/summary-statistics) and PubMed (https://pubmed.ncbi.nlm.nih.gov/) (**Supplementary table S5**). We focused on European ancestry studies with at least 2,000 study participants for which signed summary statistics were available. We chose the study with the largest cohort size, where multiple studies were available for a given trait. By applying these filters, we obtained GWAS statistics for the IgA nephropathy (IGA) (*33*), Chronic obstructive pulmonary disease (COPD) (*34*), Obesity (OB) (*34*), Psoriasis (PS) (*35*), Rheumatoid arthritis (RA) (*36*), Systemic lupus erythematosus (SLE) (*37*), Type 1 diabetes (T1D) (*38*), Thyroiditis (TRD) (*33*), Celiac disease(CeD) (*39*), Inflammatory bowel disease (IBD), which IBD summary statistics also included results for Crohn’s disease and ulcerative colitis (*40*). We have also downloaded four neurodegenerative diseases: Alzheimer’s disease (AD) (*41*), Amyotrophic lateral sclerosis (ALS) (*42*), Frontotemporal dementia (FTD) (*43*), Parkinson’s disease (PD) (*44*), four psychiatric disorders/traits: Bipolar disorder (BIP) (*45*), Major depressive disorder (MDD) (*46*), Neuroticism (Neuro) (*47*), Schizophrenia (SCZ) (*48*), and three metabolic traits: Type 2 diabetes (T2D) (*49*), Body mass index (BMI) and waist-to-hip ratio adjusted BMI (WHRadjBMI) (*50*). Given that most of the GWAS we collected were conducted in participants of European ancestry, we used the results of updated MS GWAS summary statistics in European ancestry for this analysis.

We removed the Extended MHC region (xMHC) region from the summary statistics (defined as the regions between *HIST1H2AA* and *RPL12P1* genes: chr6: 25,383,722-33,368,421Mb; ∼7.6Mb, GRCh37). We then removed indels and SNPs inconsistent with the 1000 Genomes Project (phase 3) reference panel and filtered for strand-unambiguous biallelic SNPs with minor allele frequency (MAF) >0.01 in the 1000 Genomes European (EUR) reference individuals.

### Cross-trait LD score regression

LDSC (*103*) bivariate genetic correlations attributed to genome-wide SNPs (rg) were estimated across 25 human diseases/traits from published GWASs, as mentioned above. We used LD scores from the ‘eur_w_ld_chr’ file available from https://alkesgroup.broadinstitute.org/LDSCORE, computed using 1000 Genomes Project (*93*) Europeans as a reference panel (*104*). FDR<0.05 was used to define significant genetic correlations by adjusting for the number of traits tested.

### Genome-wide polygenic score (GPS) design and optimization

We used PRS-CSx, a Bayesian polygenic modeling framework, to develop genomic prediction scores (GPS) across diverse ancestries (*23*). PRS-CSx integrates GWAS summary statistics from multiple populations, accounting for population-specific linkage disequilibrium (LD) patterns. Specifically, we utilized GWAS summary statistics from three ancestral groups: African (AFR), European (EUR), and Admixed American (AMR), and combined them using the ‘meta’ setting in PRS-CSx. In our study, 70% of the training data consisted of individuals of European ancestry from the eMERGE cohort (615 MS cases and 53,250 controls) to optimize model selection. To ensure no overlap between the GWAS discovery cohort and the GPS development dataset, the eMERGE dataset was excluded from the MS GWAS discovery cohort. We evaluated model robustness by running PRS-CSx with different values of the global shrinkage parameter: 1, 10 ¹, 10 ², 10 , 10 , and 10 . The final GPS was selected based on the best-performing model for the training dataset (**Supplementary table S9**). The score was standardized to zero mean and unit variance based on ancestry-matched population controls. In the optimization dataset, the shrinkage parameter (10^−4^) explained 2% of the variance (R2), with 1 s.d. of the score increasing MS risk by 62% (odds ratio (OR) = 1.62, 95% confidence interval (CI) = 1.49–1.75, P < 5.33 × 10^−32^) after controlling for age, sex, batch effects, and four genetic PCs. The final PRS-CSx output included 1,161,784 HapMap3 (*105*) variants and their weights.

### PheWAS

The derived polygenic predictors for MS were used to score all 102,138 eMERGE participants with available genotypes and electronic health record (EHR) data. To test the association of these polygenic predictors with diseases in a phenome-wide manner, we first harmonized the diagnostic data by converting all available ICD-10-CM codes to the ICD-9-CM system. A total of 102,138 genotyped eMERGE participants had 20,783 unique ICD-9 codes, which were subsequently mapped to 1,817 distinct phecodes. Phenome-wide association analyses (PheWAS) were conducted using the PheWAS R package (*106*), which applies predefined control groups for each phecode. For case definition, at least two occurrences of ICD-9 codes within the case grouping of each phecode were required. Logistic regression was used to test associations between the MS polygenic score and each of the 1,817 phecodes, with case-control status as the outcome. The polygenic score for MS was adjusted for age, sex, study site, and ancestry’s first three principal components (PCs). We applied a Bonferroni correction for multiple testing to determine statistically significant disease associations, setting the significance threshold at 2.75 × 10 (0.05 divided by 1,817).

### MRI analysis

Multiple sclerosis (MS) participants were from the Comprehensive Longitudinal Investigation of Multiple Sclerosis at the Brigham and Women’s Hospital (CLIMB) study (*56*). CLIMB is a natural history observational study of MS in which participants undergo semi-annual neurological examinations and annual magnetic resonance imaging (MRI). MS lesions and brain tissue compartments (gray matter, white matter, and cerebrospinal fluid) were segmented using template-driven segmentation and partial volume artifact correction (TDS+) method (*107*). Results underwent quality control and manual correction where necessary (*108*) (**Supplementary fig. S7**). MRI and genome-wide genotyping data were available for 145 MS patients; 136 of them were European ancestry, 7 were AFR ancestry, and 2 were Hispanics. Among them, 130 are diagnosed with relapsing-remitting MS, and 15 are clinically isolated syndrome. GPS score for each participant was calculated using the PLINK command ‘--bfile -- score sum –out’ (*96*), and a regression model was used to test the association between GPS and MRI, adjusted for age at visit, sex, and top three genotype PCs.

### Colocalization analysis

The COLOC package (version 5.1.0) (*109*) was applied to test the approximate Bayes factor (ABF) colocalization hypothesis, which assumes a single causal variant. Under ABF analysis, the association of a trait with a SNP is assessed by calculating the posterior probability (value from 0 to 1), with the value of 1 indicating the causal SNP. In addition, the ABF analysis has 5 hypotheses, where, PP.H0.abf indicates there is neither an eQTL nor a GWAS signal at the loci; PP.H1.abf indicates the locus is only associated with the GWAS; PP.H2.abf indicates the locus is only associated with the eQTL; PP.H3.abf indicates that both the GWAS and eQTL are associated but to a different genetic variant; PP.H4.abf indicates that the eQTL and the GWAS are associated to the same genetic variant. With the posterior probability of each SNP and aiming to find the casual variants between the GWAS and eQTL, we focused on extracting the PP.H4 value for each SNP in our study.

For MS GWAS, we used the reported lead SNPs of 236 loci. For each locus, we searched for the eSNPs that are within 500 KB of the lead SNP, and listed eGenes that were paired with the eSNP. We then obtained the eGenes cis-eQTL output around the lead eSNP within 1 Mbp window size. In addition, we extracted GWAS summary statistics around the reported 236 lead SNP. At last, we conducted COLOC for respective pair of eGene-eQTL and eSNP-GWAS for each cell type, using eQTL summary statistics from the OneK1K cohort (982 PBMC samples, 14 blood cell types, browsable results are available at www.onek1k.org) (*63*) and ROSMAP (424 DLPFC samples, 6 brain cell types, https://doi.org/10.7303/syn52335732) cohort (*62*).

### Immunohistochemistry staining for STAT3 and Glutamate decarboxylase 1 (GAD1)*+*Glutamate decarboxylase 2 (GAD2)

For validation immunostaining, a six μm formalin-fixed paraffin-embedded (FFPE) tissue section from the dorsolateral prefrontal cortex (Brodmann Area 9) of an MS individual was obtained from the New York Brain Bank at Columbia University. The tissue was stained with NeuN (1:100, 488 channel, Invitrogen, cat.# PA5-80745), STAT3 (1:100, 488 channel, Abcam cat.# ab20181), and GAD1+GAD2 (1:100, 647 channel, Wako cat.# 01919741). The FFPE tissue section was deparaffinized using CitriSolv (d-limonene, Decon Laboratories, Inc. cat.# 1601H) as a clearing agent for 20 minutes. The section was rehydrated and prepared for staining through a series of graded ethanol washes. Heat-mediated antigen retrieval was performed with citrate buffer (pH=6, Sigma-Aldrich catalog no. C9999) using a microwave (800W, 30% power setting) for 25 minutes. Following this, the section was blocked for 30 minutes at room temperature (RT) using a Bovine Serum Albumin-blocking medium (BSA, 3%, Sigma-Aldrich, catalog no. A7906) to minimize non-specific antibody binding. The section was incubated overnight with the primary antibodies (anti-STAT3 and anti-GAD1+GAD2) at 4°C. After washing, the tissues were incubated for one hour with fluorochrome-conjugated secondary antibodies (1:500, Alexa Fluor 488 and 568, Invitrogen, catalog no. A21206, A21202, A21447) to bind to the primary antibody for protein detection and signal enhancement. After washing, the slides were again incubated in 3% BSA for 30 min and stained with the NeuN-conjugated-647 antibody. After incubation, the section was washed and treated with True Black Lipofuscin Autofluorescence Quencher for 2 minutes at RT to minimize endogenous autofluorescence. An anti-fading DAPI mounting agent (347 channel, Invitrogen, catalog no. P36931) was used to coverslip.

Images were acquired using the Nikon Eclipse Ni-E immunofluorescence microscope at a magnification of ×20), and approximately 44 pictures were acquired from the MS individual. The captured images were analysed using CellProfiler (*110*) software. An extensive pipeline has been developed to automatically segment the Neurons and detect *STAT3* expressed by GAD1^+^ and GAD2^+^ cells (*111*). DAPI and NeuN was defined as the primary object using the “IdentifyPrimaryObjects” module. The Robust Background method was used for thresholding. The typical diameter for DAPI objects was set to range between 15 and 80 pixels and between 30 and 80 pixels for NEUN objects. Then, the ’RelateObjects’ module was applied to filter NEUN objects positive for DAPI objects (NEUN+DAPI+). The module “IdentifyPrimaryObjects” was used to segment GAD1/GAD2+ cells, using the Robust Background as the thresholding method, with a typical diameter ranging from 30 to 80 pixels. The segmented GAD1/GAD2+ objects were related to NEUN+DAPI+ filter GAD1/GAD2+NEUN+DAPI+ objects. The STAT3 intensity was measured within the GAD1/GAD2+NEUN+DAPI+ objects.

## Data Availability

All data produced in the present study are available upon reasonable request to the authors

## Acknowledgements

We thank the participants who donated their time, life experiences and DNA to this research and the clinical and scientific teams that worked with them.

## Author contributions

P.L.D. conceived the study and supervised the research; L.Z., A.K. performed computational analyses; M.F., F.Z., G.W., K.K., and P.L.D. provided and analyzed the data; M.T. carried out the immunofluorescence staining analysis; P.L.D. acquired the funding; L.Z., A.K., and P.L.D. wrote the original manuscript draft. L.Z., A.K., M.F., M.T., F.Z., G.W., K.K., P.L.D., and all other authors reviewed and edited the manuscript draft.

## Corresponding author

Correspondence to Philip L. De Jager.

**Appendix A: International MS Genetics Consortium collaborators**

The authors ranked alphabetically.

**The Neuro (Montreal Neurological Institute-Hospital), Montréal, QC, Canada & Department of Neurology and Neurosurgery, McGill University, Montréal, QC, Canada & Department of Human Genetics, McGill University, Montréal, QC, Canada**

Adil Harroud

**Department of Clinical Neurosciences, University of Cambridge, Cambridge, UK**

Alastair Compston, Stephen J. Sawcer

**Perron Institute for Neurological and Translational Science, University of Western Australia, Perth, Western Australia, Australia & Institute for Immunology and Infectious Diseases, Murdoch University, Perth, Western Australia, Australia**

Allan Kermode

**Department of Neurosciences, Leuven Brain Institute, KU Leuven, Leuven, Belgium**

An Goris

**Department of Neurology, School of Medicine, Technical University of Munich, Munich, Germany & Munich Cluster for Systems Neurology (SyNergy), Munich, Germany**

Bernard Hemmer

**Department of Neurology, University Hospitals Leuven, Leuven, Belgium & Department of Neurosciences, Leuven Brain Institute, KU Leuven, Leuven, Belgium**

Bénédicte Dubois

**Menzies Institute for Medical Research, University of Tasmania, Hobart, Tasmania, Australia**

Bruce Taylor

**Department of Neurology and Center of Clinical Neuroscience, First Faculty of Medicine, Charles University and General University, Prague, Czech Republic**

Dana Horakova

**Departments of Neurology and Immunobiology, Yale School of Medicine, New Haven, CT, USA**

David A. Hafler

**Department of Neurology, University General Hospital of Larissa, Faculty of Medicine, School of Health Sciences, University of Thessaly, Larissa, Greece**

Efthimios Dardiotis

D**epartment of Population and Quantitative Health Sciences, School of Medicine, Case Western Reserve University, Cleveland, OH, USA**

Farren B. S. Briggs

**Neurology Unit and Laboratory of Human Genetics of Neurological Disorders, IRCCS Ospedale San Raffaele Scientific Institute, Milan, Italy**

Federica Esposito

**Dino Ferrari Center, Department of Pathophysiology and Transplantation, University of Milan, Milan, Italy & Neurology Unit, IRCCS Fondazione Ca’ Granda Ospedale Maggiore Policlinico, Milan, Italy**

Filippo Martinelli-Boneschi

**Medical School, University of Cyprus, Nicosia, Cyprus**

Georgios Hadjigeorgiou

**Centre for Immunology and Allergy Research, The Westmead Institute for Medical Research, Westmead, New South Wales, Australia & School of Medical Sciences, Faculty of Medicine and Health, The University of Sydney, Sydney, New South Wales, Australia**

Grant P. Parnell

**Department of Neurology, Oslo University Hospital, Oslo, Norway & Institute of Clinical Medicine, University of Oslo, Oslo, Norway**

Hanne F. Harbo

**Danish Multiple Sclerosis Center, Department of Neurology, Copenhagen University Hospital– Rigshospitalet, Glostrup, Denmark**

Helle B. Søndergaard

**Department of Clinical Neuroscience, Karolinska Institutet, Center for Molecular Medicine, Karolinska University Hospital, Stockholm, Sweden**

Ingrid Kockum, Tomas Olsson

**John P. Hussman Institute for Human Genomics, Miller School of Medicine, University of Miami, Miami, FL, USA & The Dr John T. Macdonald Foundation Department of Human Genetics, Miller School of Medicine, University of Miami, Miami, FL, USA**

Jacob L. McCauley, Margaret A. Pericak-Vance

**Centre for Molecular Medicine Norway, University of Oslo, Oslo, Norway & Institute for Molecular Medicine Finland, Helsinki Institute for Life Sciences, University of Helsinki, Helsinki, Finland**

Janna Saarela

**Department of Neurology, John Hunter Hospital, Hunter New England Health District, Newcastle, New South Wales, Australia & Hunter Medical Research Institute, University of Newcastle, Newcastle, New South Wales, Australia**

Jeannette Lechner-Scott

**Departments of Neurology and Immunology, MS Center ErasMS, Erasmus University Medical Center, Rotterdam, The Netherlands & Neuroimmunology Research Group, Netherlands Institute for Neuroscience, Amsterdam, The Netherlands**

Joost Smolders

**UCSF Weill Institute for Neurosciences, Department of Neurology, University of California, San Francisco, CA, USA**

Jorge R. Oksenberg, Roland G. Henry, Sergio E. Baranzini, Stephen L. Hauser

**Department of Neurology, Johns Hopkins University School of Medicine, Baltimore, MD, USA**

Kathryn C. Fitzgerald, Peter A. Calabresi

**Genetic Epidemiology and Genomics Laboratory, Division of Epidemiology, School of Public Health, University of California, Berkeley, CA, USA**

Lisa F. Barcellos

**Servei de Neurologia-Neuroimmunologia, Centre d’Esclerosi Múltiple de Catalunya (Cemcat), Vall d’Hebron Institut de Recerca, Vall d’Hebron Hospital Universitari, Barcelona, Spain**

Manuel Comabella

**Department of Neurology, Medical University of Graz, Graz, Austria**

Michael Khalil

**Division of Psychological Medicine and Clinical Neurosciences, School of Medicine, Cardiff University, USA**

Neil Robertson

**Department of Neurology, Graduate School of Medical Sciences, Kyushu University, Fukuoka, Japan**

Noriko Isobe

**Nantes Université, CHU Nantes, Centrale Nantes, Inserm, Center for Research in Transplantation and Translational Immunology, Nantes, France**

Pierre-Antoine Gourraud, Nicolas Vince

**Institute of Experimental Immunology, University of Zurich, Zurich, Switzerland & Research and Development, Cellerys, Schlieren, Switzerland & Department of Neuroimmunology and Multiple Sclerosis Research, University Hospital Zurich, Zurich, Switzerland & Therapeutic Immune Design Unit, Department of Clinical Neuroscience, Karolinska Institutet, Stockholm, Sweden**

Roland Martin

**Department of Health Sciences and Center on Auto-immune and Allergic Diseases (CAAD), University of Eastern Piedmont, Novara, Italy**

Sandra D’alfonso

**Neurology Department, University Hospital North Midlands NHS Trust, Stoke-on-Trent, UK**

Seema Kalra

**Department of Brain and Behavioral Sciences, University of Pavia, Pavia, Italy**

Teresa Fazia

**Department of Clinical Neurosciences and the Hotchkiss Brain Institute, University of Calgary, Calgary, Alberta, Canada**

Voon Wee Yong

