## Supplemental figures and tables for "GWAS highlights the neuronal contribution to multiple sclerosis susceptibility"

Lu Zeng *et al.*

**Supplementary Figures**

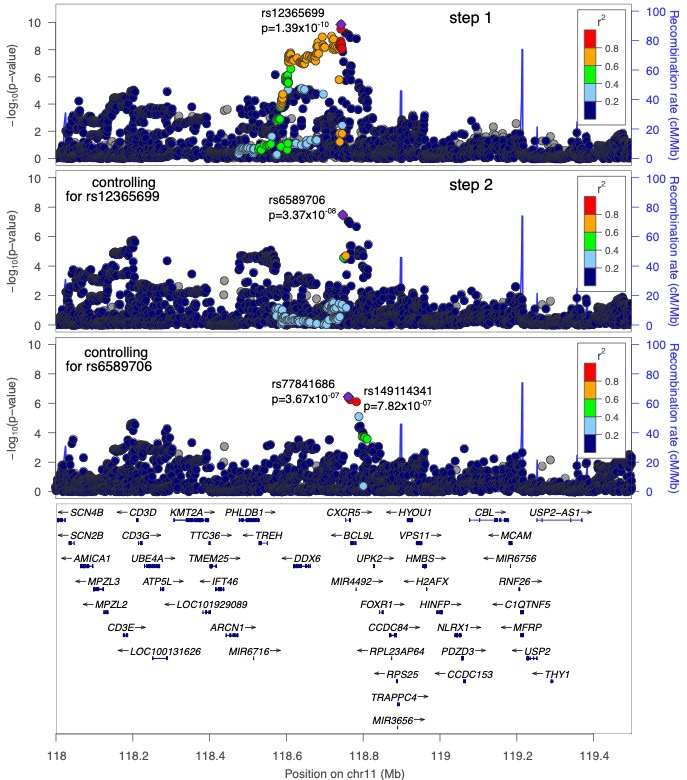

**Fig. S1. Multiple independent effects in the *DDX6* locus.** Regional association plot of the *DDX6* locus. Discovery P values (fixed-effects inverse-variance meta-analysis) are displayed. The layer tagged “Step 1” plots the associations of the marginal analysis, with the most statistically significant SNP being rs12365699 (Zscore = -4.62; P = 2.10 × 10^−19^). The “Step 2” plots the associations conditioning on rs12365699; rs6589706 is the most statistically significant SNP (Zscore = 5.67; P = 3.37 × 10^−08^). Last, “Step 3” plots the results conditioning on rs12365699 and rs6589706, with rs77841686 displaying the lowest P value (Zscore = -6.52; P = 3.67 × 10^−07^), which is LD linked (R2=0.96) with rs149114341 (Zscore = -6.05; P = 7.82 × 10^−07^). All three SNPs reached genome-wide significance in the meta-analysis of European Ancestry. Each of the three independent SNPs—lead SNPs—are highlighted by use of a triangle in the respective layer.

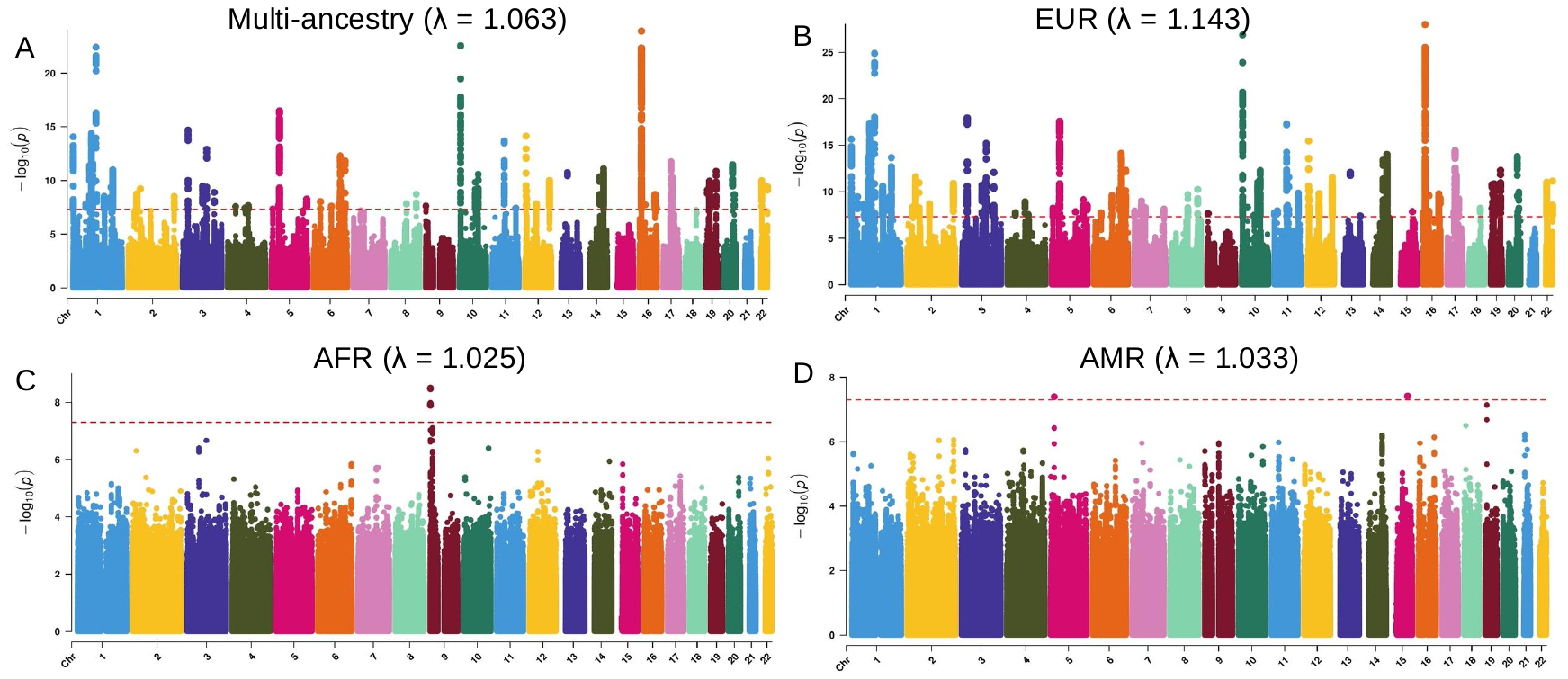

**Fig. S2. Manhattan plot of genome-wide association meta-analysis of multi-ancestry (A), European ancestry (EUR) (B), African American (AFR) (C) and Admixed American (AMR) (D).** The x axis shows genomic position (chromosomes 1–22), and the y axis shows statistical significance as –log10(P-value). P values are two-sided and based on an inverse-variance-weighted fixed-effects meta-analysis. The red line shows the genome-wide significance threshold (P < 5 × 10^−8^). The genomic inflation factor (λ) is highlighted in each cohort.

**Fig. S3.** **Selected markers and top differentially expressed genes between subpopulations.** Gene expression (rows) across subpopulations (columns) of astrocytes, excitatory neurons, inhibitory neurons, microglia, oligodendrocytes and OPCs. Dot color: mean expression in expressing cells. Dot size: percent of cells expressing the gene.

A

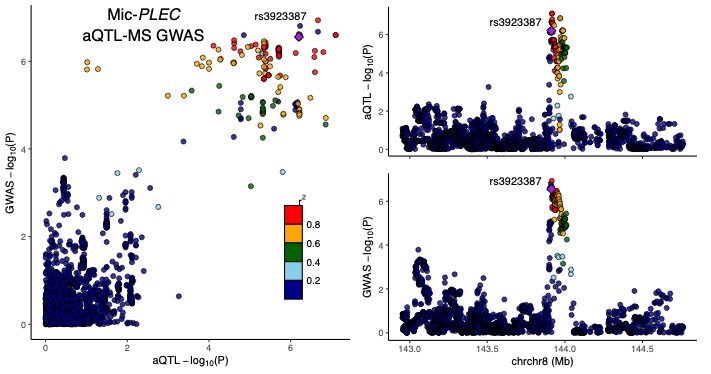

B

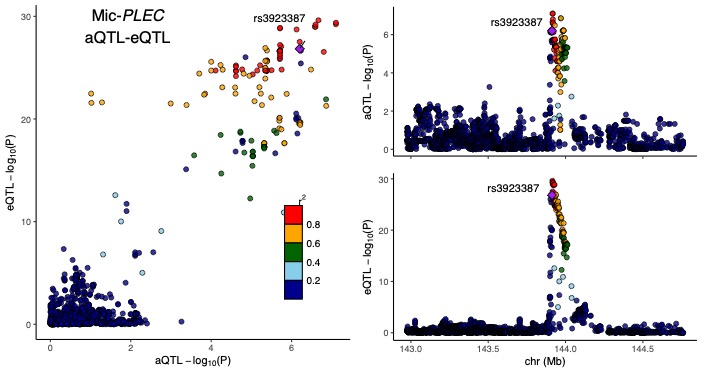

**Fig. S4. Example of MS GWAS signals and eQTL signals compared to ATACseq-QTLs. A) and B).** Locuszoom plot around the *PLEC* locus in microglia. The y-axis shows -log10 of p-values for the MS GWAS/eQTL and x-axis shows -log10 of p-values for the ATAC-seq aQTL in corresponding cell type.

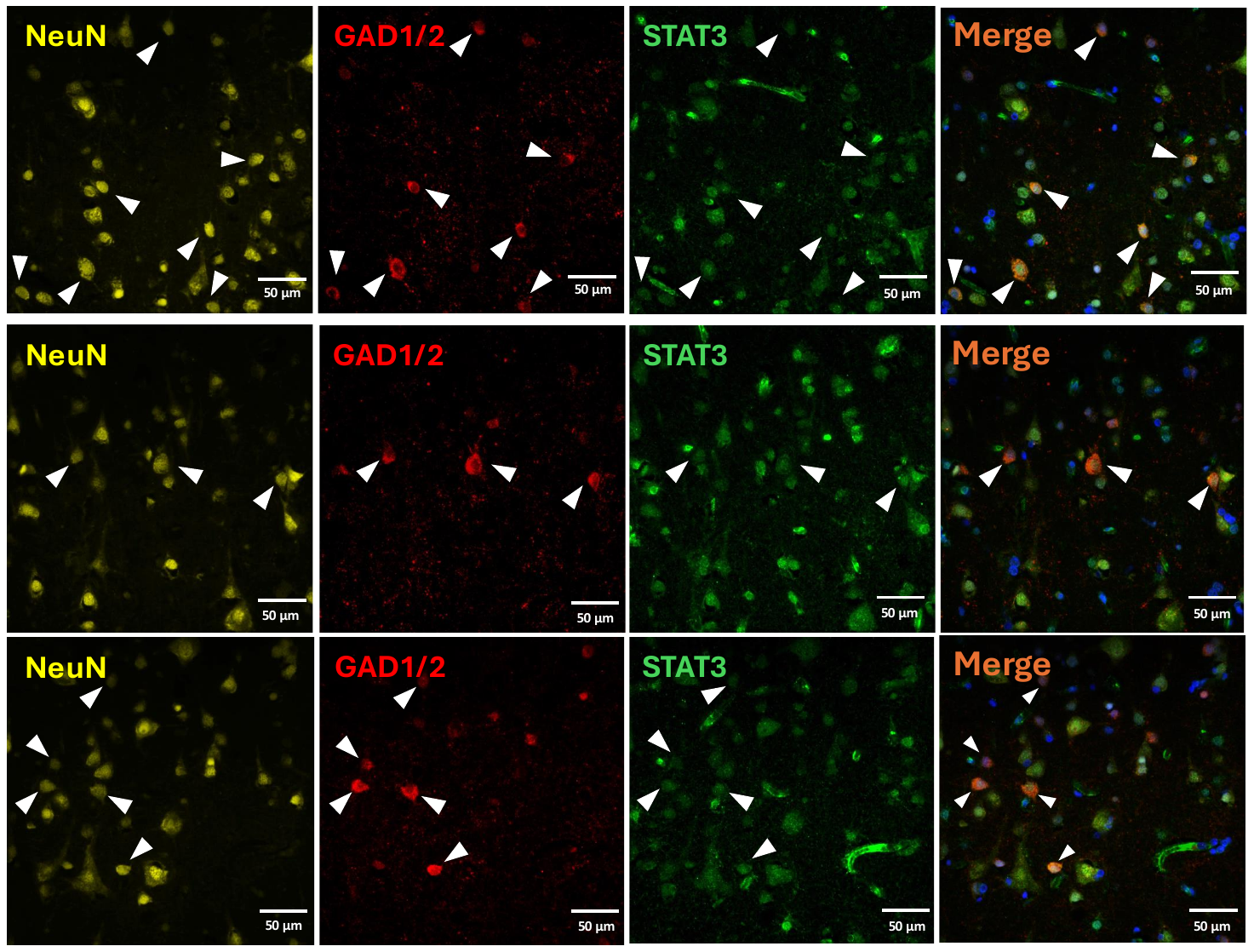

**Fig. S5.** Immunohistochemistry of DLPFC in human MS brain tissue, stained for STAT3 (green), GAD1/2 (red), and NeuN (yellow), with DAPI (blue) to visualize nuclei. Expression of STAT3 was observed in NeuN+GAD1/2+ neurons. White triangles highlight the colocalization of DAPI, STAT3, GAD1/2, and NeuN. Scale bar, = 50 μm.

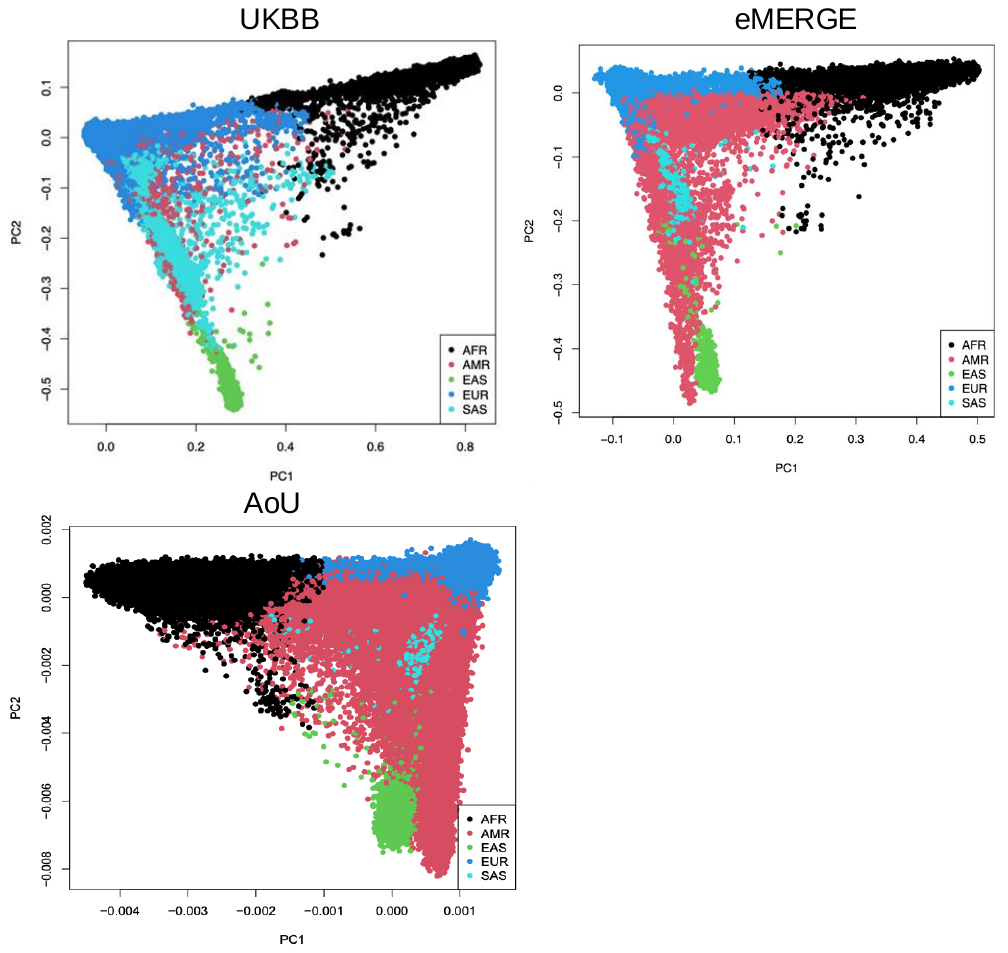

**Fig. S6.** 3 PCA projections of the study participants from the UKBB (top), eMERGE-III (right) and AoU (bottom) against the 1000 G reference populations.

**Fig. S7.** MRI data distribution of brain parenchymal fraction (tds_ bpf), lesion volume (tds_ lesion), total intracranial volume (tds_ icc), total white matter volume (tds_ wm), total gray matter volume (tds_ gm) and total cerebrospinal fluid volume (tds_csf) in log transformed and non-log (top) transformed (bottom).

**Supplementary Tables**

**Table S1. Descriptive statistics of the new GWAS data of the discovery phase.**

|  | **UKBB** | | **eMERGE** | | **AoU** | |
| --- | --- | --- | --- | --- | --- | --- |
| **EUR** | Case (n=2259) | Control (n=390969) | Case (n=902) | Control (n=76164) | Case (n=1902) | Control (n=129207) |
| **Age** | 55.397 | 57.003 | 63.977 | 60.750 | 58.103 | 60.179 |
| **Females:Males** | 1537:720 | 212094:178846 | 649:253 | 39491:36672 | 1395:456 | 75657:50072 |
| **AFR** |  |  | Case (n=143) | Control (n=15719) | Case (n=471) | Control (n=46325) |
| **Age** |  |  | 58.853 | 47.782 | 54.236 | 54.093 |
| **Females:Males** |  |  | 116:27 | 9439:6280 | 378:83 | 27641:17496 |
| **AMR** |  |  | Case (n=55) | Control (n=5168) | Case (n=297) | Control (n=38965) |
| **Age** |  |  | 60.855 | 56.406 | 48.625 | 48.871 |
| **Females:Males** |  |  | 37:18 | 3041:2127 | 232:56 | 26139:12045 |

**Table S2. The list of 236 independent MS risk variants in European ancestry.**

The prior p-value refers to the p-value reported in the 2019 IMSGC paper, while the current p-value is derived from the meta-analysis (METAL) conducted in this study.

| **Locus** | **CHR** | **BP (GRCh37)** | **Lead variant** | **Prior *P-*value/**  **Current P-value** | **Beta (SE)** | **EA/OA** | **Novel/**  **Known** |
| --- | --- | --- | --- | --- | --- | --- | --- |
| 1 | 1 | 2520527 | rs6670198 | 1.54E-36/2.50E-15 | 0.049 (0.006) | T/C | Known |
| 2 | 1 | 2701575 | rs375915427 | 3.15E-08 | 0.076 (0.014) | T/C | Novel |
| 3 | 1 | 6512547 | rs2986736 | 8.91E-17/2.21E-06 | -0.034 (0.007) | T/C | Known |
| 4 | 1 | 11883342 | rs198398 | 2.27E-09/5.17E-05 | -0.053 (0.013) | T/C | Known |
| 5 | 1 | 24207504 | rs67934705 | 4.36E-08/1.01E-05 | -0.033 (0.007) | A/G | Known |
| 6 | 1 | 25291010 | rs6672420 | 1.48E-09/2.22E-06 | 0.028 (0.006) | A/T | Known |
| 7 | 1 | 32738415 | rs79979643 | 1.44E-15/6.54E-09 | 0.047 (0.008) | A/G | Known |
| 8 | 1 | 65429319 | rs72922276 | 1.41E-15/3.39E-09 | -0.060 (0.010) | A/G | Known |
| 9 | 1 | 85682020 | rs11161550 | 2.06E-09/0.006824 | -0.016 (0.006) | A/G | Known |
| 10 | 1 | 85729820 | rs35486093 | 2.27E-31/3.19E-14 | -0.079 (0.010) | A/G | Known |
| 11 | 1 | 85748811 | rs529392609 | 2.63E-10 | -0.724 (0.115) | A/G | Novel |
| 12 | 1 | 85764886 | rs11161589 | 6.12E-12 | -0.041 (0.006) | A/G | Novel |
| 13 | 1 | 92222089 | rs12133753 | 1.67E-16/3.68E-10 | -0.051 (0.008) | T/C | Known |
| 14 | 1 | 92939959 | rs58394161 | 1.37E-14/3.17E-15 | -0.059 (0.008) | T/C | Known |
| 15 | 1 | 92956978 | rs113561235 | 3.60E-08 | 0.097 (0.018) | T/C | Novel |
| 16 | 1 | 92973242 | rs79285232 | 5.85E-11 | -0.184 (0.028) | T/C | Novel |
| 17 | 1 | 93088923 | rs72724541 | 1.26E-11 | 0.127 (0.019) | A/G | Novel |
| 18 | 1 | 93152635 | rs11809700 | 2.95E-30/6.75E-18 | 0.056 (0.006) | T/C | Known |
| 19 | 1 | 93291944 | rs12042488 | 1.09E-11 | 0.062 (0.009) | A/T | Novel |
| 20 | 1 | 93426869 | rs1415069 | 3.32E-08/0.00298 | -0.023 (0.008) | C/G | Known |
| 21 | 1 | 101289496 | rs142860878 | 1.77E-08 | -0.181 (0.032) | C/G | Novel |
| 22 | 1 | 101290432 | rs34723276 | 1.11E-22/5.55E-15 | 0.052 (0.007) | A/G | Known |
| 23 | 1 | 101307053 | rs12047318 | 4.93E-10 | -0.068 (0.011) | A/G | Novel |
| 24 | 1 | 101412902 | rs11578655 | 2.89E-08/6.97E-08 | -0.046 (0.008) | T/G | Known |
| 25 | 1 | 101544143 | rs147885102 | 3.69E-10 | -0.050 (0.008) | A/T | Novel |
| 26 | 1 | 117090493 | rs10801908 | 4.55E-70/1.45E-24 | -0.087 (0.008) | T/C | Known |
| 27 | 1 | 120267505 | rs483180 | 1.45E-17/1.25E-10 | 0.041 (0.006) | C/G | Known |
| 28 | 1 | 154983036 | rs112344141 | 1.67E-08/0.000208 | -0.056 (0.015) | T/G | Known |
| 29 | 1 | 157660829 | rs77191363 | 7.86E-09 | 0.075 (0.013) | C/G | Novel |
| 30 | 1 | 157686337 | rs2317231 | 3.25E-16/2.70E-11 | -0.039 (0.006) | T/G | Known |
| 31 | 1 | 160389984 | rs3737798 | 4.12E-14/2.41E-05 | 0.025 (0.006) | A/G | Known |
| 32 | 1 | 160634588 | rs6427540 | 1.48E-09/6.45E-09 | -0.049 (0.008) | T/C | Known |
| 33 | 1 | 160703965 | rs983494 | 6.10E-16/2.57E-07 | -0.037 (0.007) | A/G | Known |
| 34 | 1 | 192541021 | rs1323292 | 3.64E-33/3.63E-10 | 0.047 (0.008) | A/G | Known |
| 35 | 1 | 200875897 | rs59655222 | 5.66E-21/3.18E-13 | 0.048 (0.007) | T/C | Known |
| 36 | 1 | 212877776 | rs9308424 | 9.25E-14/1.18E-07 | -0.033 (0.006) | A/G | Known |
| 37 | 2 | 12607893 | rs11899404 | 1.66E-08/0.0003779 | -0.021 (0.006) | T/C | Known |
| 38 | 2 | 25052177 | rs11125803 | 5.45E-10/1.18E-05 | -0.030 (0.007) | T/C | Known |
| 39 | 2 | 30472442 | rs13414105 | 6.90E-10/9.40E-06 | -0.044 (0.010) | A/C | Known |
| 40 | 2 | 30478386 | rs4952115 | 4.31E-09 | -0.046 (0.008) | T/G | Novel |
| 41 | 2 | 43355324 | rs12478539 | 1.65E-24/9.19E-12 | -0.045 (0.007) | C/G | Known |
| 42 | 2 | 61066666 | rs1432295 | 2.74E-08 | -0.033 (0.006) | A/G | Novel |
| 43 | 2 | 61242410 | rs1177228 | 2.31E-19/1.63E-06 | -0.033 (0.007) | A/G | Known |
| 44 | 2 | 65661843 | rs13385171 | 3.47E-11/8.84E-08 | -0.032 (0.006) | T/C | Known |
| 45 | 2 | 68646536 | rs12622670 | 5.08E-16/8.44E-07 | 0.029 (0.006) | T/C | Known |
| 46 | 2 | 112492986 | rs71252597 | 1.78E-12/2.02E-09 | -0.043 (0.007) | T/C | Known |
| 47 | 2 | 112770799 | rs57116599 | 1.11E-11/1.89E-07 | -0.036 (0.007) | A/G | Known |
| 48 | 2 | 136884679 | rs10191360 | 3.85E-10/0.0004101 | 0.021 (0.006) | T/C | Known |
| 49 | 2 | 151644203 | rs962052 | 8.28E-09/0.0001346 | -0.024 (0.006) | T/C | Known |
| 50 | 2 | 191989356 | rs6738544 | 2.40E-13/3.97E-06 | -0.029 (0.006) | A/C | Known |
| 51 | 2 | 204632861 | rs12614091 | 6.18E-12/0.0001717 | 0.026 (0.007) | A/T | Known |
| 52 | 2 | 231121829 | rs35540610 | 2.98E-33/8.78E-11 | -0.047 (0.007) | T/C | Known |
| 53 | 3 | 18798848 | rs9863496 | 2.75E-14/0.0008083 | -0.023 (0.007) | T/C | Known |
| 54 | 3 | 27783015 | rs13327021 | 1.55E-16/2.41E-10 | 0.038 (0.006) | T/C | Known |
| 55 | 3 | 28072086 | rs438613 | 2.31E-49/1.15E-18 | -0.052 (0.006) | T/C | Known |
| 56 | 3 | 32962051 | rs11919880 | 7.63E-10/1.42E-06 | 0.029 (0.006) | A/G | Known |
| 57 | 3 | 71535338 | rs9878602 | 1.91E-18/1.37E-06 | 0.028 (0.006) | T/G | Known |
| 58 | 3 | 100848597 | rs111430408 | 6.42E-11/5.77E-06 | -0.048 (0.011) | T/C | Known |
| 59 | 3 | 101661456 | rs74482986 | 2.46E-08 | -0.063 (0.011) | A/C | Novel |
| 60 | 3 | 101749022 | rs4325907 | 1.99E-09/1.98E-11 | -0.041 (0.006) | T/C | Known |
| 61 | 3 | 105455955 | rs2289746 | 4.59E-12/7.67E-08 | -0.033 (0.006) | T/C | Known |
| 62 | 3 | 112693983 | rs138433213 | 3.24E-09/0.0003066 | 0.094 (0.026) | T/G | Known |
| 63 | 3 | 119228508 | rs9843355 | 4.14E-30/6.93E-12 | -0.054 (0.008) | A/G | Known |
| 64 | 3 | 121542898 | rs2331964 | 4.91E-20/1.08E-09 | -0.037 (0.006) | T/C | Known |
| 65 | 3 | 121765368 | rs71329256 | 6.98E-43 |  |  | Known |
| 66 | 3 | 121770539 | rs2255214 | 6.19E-16 | -0.047 (0.006) | T/G | Novel |
| 67 | 3 | 121783015 | rs75937181 | 8.32E-10/3.44E-11 | 0.068 (0.010) | T/G | Known |
| 68 | 3 | 141150990 | rs6789653 | 1.30E-09/0.001328 | -0.020 (0.006) | A/G | Known |
| 69 | 3 | 159691112 | rs1014486 | 3.13E-28/8.46E-13 | -0.042 (0.006) | T/C | Known |
| 70 | 3 | 159702290 | rs9858816 | 2.64E-08 | -0.034 (0.006) | T/C | Novel |
| 71 | 3 | 159712373 | rs10936182 | 4.99E-08 |  |  | Known |
| 72 | 3 | 169536637 | rs10936602 | 1.90E-11/9.75E-07 | 0.033 (0.007) | T/C | Known |
| 73 | 3 | 187565968 | rs2590438 | 4.65E-09/2.33E-05 | -0.026 (0.006) | T/G | Known |
| 74 | 3 | 187987624 | rs13066789 | 1.50E-08/0.001354 | -0.019 (0.006) | T/C | Known |
| 75 | 4 | 40307564 | rs13136820 | 7.79E-10/2.42E-08 | -0.035 (0.006) | T/C | Known |
| 76 | 4 | 48127262 | rs6837324 | 1.34E-11/1.89E-05 | -0.026 (0.006) | A/G | Known |
| 77 | 4 | 87862396 | rs2705616 | 1.15E-11/1.18E-09 | 0.036 (0.006) | C/G | Known |
| 78 | 4 | 103911781 | rs6533052 | 7.03E-16/4.55E-06 | 0.027 (0.006) | A/G | Known |
| 79 | 4 | 106255589 | rs2726479 | 3.84E-11/1.70E-05 | -0.026 (0.006) | T/C | Known |
| 80 | 4 | 109058718 | rs9992763 | 2.36E-11/1.01E-07 | -0.031 (0.006) | T/G | Known |
| 81 | 4 | 122119449 | rs17051321 | 1.41E-10/0.0001026 | 0.027 (0.007) | T/C | Known |
| 82 | 4 | 164493807 | rs72989863 | 5.55E-09/9.07E-05 | -0.024 (0.006) | A/G | Known |
| 83 | 5 | 6712834 | rs34681760 | 2.01E-11/6.96E-09 | -0.036 (0.006) | T/C | Known |
| 84 | 5 | 35877505 | rs10063294 | 1.58E-28/2.50E-09 | -0.035 (0.006) | A/G | Known |
| 85 | 5 | 40393852 | rs1992662 | 1.67E-17 | 0.052 (0.006) | A/G | Novel |
| 86 | 5 | 40396425 | rs11749040 | 4.54E-25/2.61E-18 | 0.075 (0.009) | A/G | Known |
| 87 | 5 | 40429250 | rs2889382 | 1.02E-11/0.01198 | -0.015 (0.006) | A/T | Known |
| 88 | 5 | 55444683 | rs7731626 | 3.89E-15/6.07E-08 | -0.033 (0.006) | A/G | Known |
| 89 | 5 | 118703662 | rs32658 | 3.24E-08/3.58E-05 | 0.025 (0.006) | T/G | Known |
| 90 | 5 | 118815815 | rs28762138 | 1.46E-08 | -0.219 (0.039) | T/G | Novel |
| 91 | 5 | 133449827 | rs244656 | 2.96E-14/1.06E-07 | 0.043 (0.008) | A/T | Known |
| 92 | 5 | 133891282 | rs2084007 | 1.91E-13/1.71E-06 | -0.028 (0.006) | T/C | Known |
| 93 | 5 | 141539339 | rs249677 | 5.73E-13/1.33E-05 | 0.027 (0.006) | A/C | Known |
| 94 | 5 | 158759900 | rs2546890 | 5.30E-19/7.48E-10 | 0.036 (0.006) | A/G | Known |
| 95 | 5 | 158944266 | rs7727104 | 8.17E-09 | 0.038 (0.007) | A/G | Novel |
| 96 | 5 | 176790162 | rs67111717 | 3.18E-21/3.62E-09 | -0.036 (0.006) | A/G | Known |
| 97 | 6 | 7100029 | rs12211604 | 2.15E-08/1.60E-07 | -0.032 (0.006) | A/G | Known |
| 98 | 6 | 14691215 | rs111635774 | 1.38E-17/3.19E-05 | -0.042 (0.010) | T/C | Known |
| 99 | 6 | 16672760 | rs719316 | 1.62E-13/0.001555 | 0.019 (0.006) | T/C | Known |
| 100 | 6 | 36348689 | rs1076928 | 2.75E-19/9.78E-08 | 0.031 (0.006) | T/C | Known |
| 101 | 6 | 90976768 | rs72928038 | 8.38E-29/2.53E-09 | 0.045 (0.008) | A/G | Known |
| 102 | 6 | 119215402 | rs11542663 | 3.24E-10/5.08E-07 | 0.030 (0.006) | A/C | Known |
| 103 | 6 | 128280104 | rs802730 | 1.08E-09/5.34E-11 | 0.043 (0.006) | T/C | Known |
| 104 | 6 | 130348257 | rs75191738 | 2.59E-06/0.00125 | 0.027 (0.008) | T/C | Known |
| 105 | 6 | 135495226 | rs2327586 | 9.48E-20/1.19E-08 | 0.038 (0.007) | T/C | Known |
| 106 | 6 | 135749682 | rs13218824 | 4.01E-08 | -0.076 (0.014) | T/C | Novel |
| 107 | 6 | 135833463 | rs4896153 | 2.72E-29/8.49E-15 | -0.047 (0.006) | A/T | Known |
| 108 | 6 | 135904197 | rs76892387 | 1.44E-08 | -0.081 (0.014) | A/G | Novel |
| 109 | 6 | 137438057 | rs62420820 | 9.26E-36/7.36E-12 | 0.048 (0.007) | A/G | Known |
| 110 | 6 | 137959455 | rs631204 | 4.92E-25/4.79E-14 | 0.045 (0.006) | A/C | Known |
| 111 | 6 | 138179146 | rs17780048 | 5.15E-12/1.07E-05 | -0.057 (0.013) | T/C | Known |
| 112 | 6 | 143865221 | rs6911131 | 1.31E-10/2.23E-05 | -0.053 (0.013) | A/G | Known |
| 113 | 6 | 159465977 | rs1738074 | 3.48E-35/5.22E-13 | -0.043 (0.006) | T/C | Known |
| 114 | 7 | 2443302 | rs55858457 | 2.95E-08/1.84E-06 | 0.030 (0.006) | T/G | Known |
| 115 | 7 | 3139417 | rs10951042 | 1.50E-18/2.41E-08 | -0.034 (0.006) | T/C | Known |
| 116 | 7 | 27135314 | rs10951154 | 3.05E-10/0.08101 | -0.014 (0.008) | T/C | Known |
| 117 | 7 | 28142186 | rs10245867 | 1.22E-11/7.99E-05 | 0.025 (0.006) | T/G | Known |
| 118 | 7 | 37382465 | rs60600003 | 4.60E-19/9.78E-10 | -0.060 (0.010) | T/G | Known |
| 119 | 7 | 50239880 | rs10230723 | 7.69E-11/3.69E-06 | 0.037 (0.008) | A/T | Known |
| 120 | 7 | 50328339 | rs116877451 | 2.58E-16/4.17E-08 | 0.091 (0.017) | A/G | Known |
| 121 | 7 | 56091706 | rs6975311 | 5.30E-09 | -0.038 (0.007) | A/G | Novel |
| 122 | 7 | 105706462 | rs73414214 | 6.34E-10/6.80E-05 | -0.038 (0.009) | A/C | Known |
| 123 | 7 | 128573967 | rs4728142 | 3.37E-09/3.99E-05 | 0.024 (0.006) | A/G | Known |
| 124 | 7 | 138729795 | rs10271373 | 3.11E-09/1.04E-05 | 0.026 (0.006) | A/C | Known |
| 125 | 7 | 149289464 | rs354033 | 7.99E-10/9.87E-09 | -0.038 (0.007) | A/G | Known |
| 126 | 8 | 79417222 | rs28703878 | 5.27E-22/2.07E-10 | -0.040 (0.006) | A/G | Known |
| 127 | 8 | 95851818 | rs78727559 | 3.23E-10/0.0002081 | -0.057 (0.015) | T/G | Known |
| 128 | 8 | 128175696 | rs735542 | 5.66E-13/2.62E-06 | 0.029 (0.006) | A/G | Known |
| 129 | 8 | 128814091 | rs6990534 | 5.85E-20/5.60E-11 | -0.041 (0.006) | A/G | Known |
| 130 | 8 | 129177769 | rs7819665 | 2.62E-25/4.61E-07 | -0.036 (0.007) | T/C | Known |
| 131 | 8 | 144986793 | rs3923387 | 1.76E-08/2.78E-07 | 0.030 (0.006) | T/C | Known |
| 132 | 9 | 4981602 | rs10758669 | 2.20E-08 | -0.034 (0.006) | A/C | Novel |
| 133 | 9 | 100868189 | rs7855251 | 1.23E-10/1.97E-06 | 0.031 (0.006) | T/C | Known |
| 134 | 10 | 6070273 | rs12722559 | 1.76E-15/1.97E-21 | -0.081 (0.009) | A/C | Known |
| 135 | 10 | 8098719 | rs1399180 | 7.16E-11/1.14E-05 | -0.035 (0.008) | T/C | Known |
| 136 | 10 | 31395761 | rs1087056 | 3.50E-19/1.30E-07 | 0.031 (0.006) | A/G | Known |
| 137 | 10 | 64384640 | rs77051803 | 3.30E-08 | 0.052 (0.009) | A/G | Known |
| 138 | 10 | 64449549 | rs61863928 | 3.01E-16/3.84E-05 | -0.025 (0.006) | T/G | Known |
| 139 | 10 | 75653800 | rs17741873 | 4.69E-08/0.00146 | -0.024 (0.007) | T/G | Known |
| 140 | 10 | 81059335 | rs1250551 | 1.68E-23/1.51E-10 | 0.041 (0.006) | T/G | Known |
| 141 | 10 | 94479107 | rs1112718 | 2.08E-17/5.36E-13 | 0.043 (0.006) | A/G | Known |
| 142 | 11 | 321138 | rs35218683 | 1.36E-08 |  |  | Known |
| 143 | 11 | 321235 | rs56232455 | 1.78E-08 | 0.041 (0.007) | A/G | Novel |
| 144 | 11 | 14402930 | rs61884005 | 6.34E-09/1.18E-06 | 0.042 (0.009) | C/G | Known |
| 145 | 11 | 14868316 | rs570429157 | 8.76E-12/2.26E-07 | -0.141 (0.027) | A/G | Known |
| 146 | 11 | 36438075 | rs1365120 | 4.06E-08/1.59E-06 | -0.048 (0.010) | T/C | Known |
| 147 | 11 | 47360412 | rs2269434 | 1.47E-13/2.79E-06 | -0.029 (0.006) | T/C | Known |
| 148 | 11 | 60783062 | rs75064517 | 6.85E-09 | -0.115 (0.020) | A/G | Novel |
| 149 | 11 | 60793651 | rs4939490 | 2.00E-29/5.62E-18 | -0.051 (0.006) | C/G | Known |
| 150 | 11 | 60827933 | rs11230581 | 5.55E-15 | 0.046 (0.006) | T/C | Novel |
| 151 | 11 | 64095178 | rs11231749 | 1.12E-11/3.02E-05 | -0.027 (0.006) | T/C | Known |
| 152 | 11 | 65705432 | rs531612 | 1.21E-09/1.20E-08 | 0.033 (0.006) | T/C | Known |
| 153 | 11 | 72450091 | rs77267834 | 2.71E-12 | 0.098 (0.014) | A/T | Novel |
| 154 | 11 | 95311422 | rs4409785 | 6.87E-12/1.45E-06 | -0.036 (0.007) | T/C | Known |
| 155 | 11 | 95421830 | rs56095240 | 1.13E-09/0.000154 | 0.031 (0.008) | A/T | Known |
| 156 | 11 | 118480695 | rs34026809 | 7.68E-17/0.0002224 | -0.042 (0.011) | C/G | Known |
| 157 | 11 | 118743286 | rs12365699 | 2.10E-19/1.39E-10 | -0.051 (0.008) | A/G | Known |
| 158 | 11 | 118747813 | rs6589706 | 4.53E-26/1.40E-08 | 0.033 (0.006) | A/G | Known |
| 159 | 11 | 118783424 | rs149114341 | 8.76E-11/1.41E-09 | -0.119 (0.020) | A/G | Known |
| 160 | 11 | 122518525 | rs6589939 | 1.75E-12/0.0001632 | -0.023 (0.006) | A/G | Known |
| 161 | 11 | 128421175 | rs4262739 | 4.41E-12/1.16E-06 | -0.029 (0.006) | A/G | Known |
| 162 | 12 | 6440009 | rs1800693 | 2.24E-47/3.54E-16 | -0.048 (0.006) | T/C | Known |
| 163 | 12 | 6441622 | rs12832171 | 4.13E-10 |  |  | Known |
| 164 | 12 | 6514963 | rs2364485 | 1.51E-20/5.78E-06 | 0.042 (0.009) | A/C | Known |
| 165 | 12 | 9866349 | rs7977720 | 4.96E-24/8.52E-11 | 0.038 (0.006) | T/C | Known |
| 166 | 12 | 58106836 | rs701006 | 9.63E-31/1.58E-10 | -0.038 (0.006) | A/G | Known |
| 167 | 12 | 94661453 | rs61708525 | 1.80E-08/0.0005186 | -0.022 (0.006) | A/G | Known |
| 168 | 12 | 111884608 | rs3184504 | 4.24E-11/5.37E-06 | 0.027 (0.006) | T/C | Known |
| 169 | 12 | 123604053 | rs7975763 | 2.99E-13/2.82E-12 | 0.050 (0.007) | T/C | Known |
| 170 | 13 | 50811220 | rs9591325 | 1.26E-19/8.34E-13 | 0.092 (0.013) | T/C | Known |
| 171 | 13 | 50961957 | rs9568402 | 4.80E-09/0.003955 | -0.026 (0.009) | A/T | Known |
| 172 | 13 | 100026952 | rs77654077 | 8.98E-17/2.90E-07 | 0.095 (0.018) | A/C | Known |
| 173 | 14 | 52306091 | rs11852059 | 1.19E-09/2.28E-07 | -0.038 (0.007) | A/C | Known |
| 174 | 14 | 69253364 | rs12434551 | 4.14E-17/4.12E-14 | 0.044 (0.006) | A/T | Known |
| 175 | 14 | 76014298 | rs34695601 | 5.56E-13/4.53E-07 | 0.034 (0.007) | T/C | Known |
| 176 | 14 | 88407917 | rs12432149 | 4.08E-12 | 0.041 (0.006) | A/G | Novel |
| 177 | 14 | 88523488 | rs116899835 | 1.67E-30/7.78E-14 | -0.101 (0.013) | T/C | Known |
| 178 | 14 | 103230758 | rs12588969 | 1.82E-09/9.49E-05 | -0.025 (0.006) | C/G | Known |
| 179 | 14 | 103265844 | rs12147246 | 1.98E-15/1.56E-07 | 0.032 (0.006) | A/G | Known |
| 180 | 15 | 79247482 | rs62013236 | 8.53E-17/3.69E-07 | -0.042 (0.008) | T/C | Known |
| 181 | 15 | 90887584 | rs6496663 | 1.13E-12/2.17E-07 | -0.033 (0.006) | A/C | Known |
| 182 | 16 | 1067832 | rs405343 | 3.05E-15/7.31E-08 | 0.041 (0.008) | T/G | Known |
| 183 | 16 | 11053656 | rs117283010 | 3.07E-16 | 0.123 (0.015) | A/G | Novel |
| 184 | 16 | 11114512 | rs2286974 | 4.09E-09/2.90E-26 | 0.063 (0.006) | A/G | Known |
| 185 | 16 | 11185464 | rs55898143 | 1.38E-13 | 0.078 (0.011) | T/C | Novel |
| 186 | 16 | 11242497 | rs794423 | 1.62E-10 | 0.099 (0.015) | A/C | Novel |
| 187 | 16 | 11247847 | rs80207443 | 1.60E-13 | 0.101 (0.014) | T/C | Novel |
| 188 | 16 | 11335999 | rs814260 | 9.30E-09 | -0.035 (0.006) | A/G | Novel |
| 189 | 16 | 11353879 | rs146566517 | 5.01E-16/4.79E-07 | 0.084 (0.017) | T/C | Known |
| 190 | 16 | 11398467 | rs10852332 | 4.02E-09 | 0.042 (0.007) | C/G | Novel |
| 191 | 16 | 11412926 | rs34947566 | 1.30E-23/1.30E-09 | -0.049 (0.008) | A/C | Known |
| 192 | 16 | 30103160 | rs3809627 | 1.23E-21/2.60E-10 | -0.038 (0.006) | A/C | Known |
| 193 | 16 | 57077094 | rs8062446 | 4.30E-09/2.25E-07 | 0.031 (0.006) | T/C | Known |
| 194 | 16 | 79111297 | rs12925972 | 1.11E-19/1.69E-10 | -0.038 (0.006) | T/C | Known |
| 195 | 16 | 79350204 | rs17724508 | 6.07E-14/4.63E-07 | 0.075 (0.015) | T/C | Known |
| 196 | 16 | 79652720 | rs6564681 | 3.70E-14/7.97E-06 | -0.028 (0.006) | T/C | Known |
| 197 | 16 | 86021505 | rs35703946 | 2.83E-17/5.00E-10 | -0.053 (0.009) | A/G | Known |
| 198 | 17 | 34842521 | rs4796224 | 1.13E-15/2.77E-08 | -0.033 (0.006) | A/G | Known |
| 199 | 17 | 37970149 | rs9909593 | 8.57E-17/5.69E-05 | -0.024 (0.006) | A/G | Known |
| 200 | 17 | 38252660 | rs883871 | 8.56E-14/4.48E-06 | 0.038 (0.008) | A/G | Known |
| 201 | 17 | 40508559 | rs58905292 | 1.96E-09 | 0.101 (0.017) | A/T | Novel |
| 202 | 17 | 40529835 | rs1026916 | 2.32E-28/5.00E-15 | 0.048 (0.006) | A/G | Known |
| 203 | 17 | 43407670 | rs7222450 | 2.09E-10/5.91E-06 | 0.027 (0.006) | A/G | Known |
| 204 | 17 | 45702280 | rs11079784 | 1.99E-27/1.64E-12 | -0.041 (0.006) | T/C | Known |
| 205 | 17 | 57859210 | rs2150879 | 4.05E-31/7.14E-10 | -0.036 (0.006) | A/G | Known |
| 206 | 17 | 57963873 | rs1292052 | 1.49E-09 | -0.070 (0.012) | T/C | Novel |
| 207 | 17 | 73335776 | rs9900529 | 4.76E-08/9.43E-07 | 0.033 (0.007) | C/G | Known |
| 208 | 18 | 56269737 | rs4940730 | 3.90E-10/0.01111 | 0.015 (0.006) | A/G | Known |
| 209 | 18 | 56348044 | rs9955954 | 4.85E-11/5.96E-09 | 0.041 (0.007) | A/G | Known |
| 210 | 18 | 67544046 | rs2469434 | 2.92E-08/0.0004944 | -0.021 (0.006) | T/C | Known |
| 211 | 19 | 4466466 | rs12971909 | 5.56E-09/8.98E-06 | 0.033 (0.007) | A/G | Known |
| 212 | 19 | 6668972 | rs1077667 | 7.88E-33/2.14E-11 | -0.047 (0.007) | T/C | Known |
| 213 | 19 | 10463118 | rs34536443 | 3.39E-11/1.98E-10 | -0.112 (0.018) | C/G | Known |
| 214 | 19 | 10592144 | rs28834106 | 3.61E-19/1.53E-10 | 0.044 (0.007) | T/C | Known |
| 215 | 19 | 11173928 | rs12609500 | 5.27E-09/1.08E-05 | -0.030 (0.007) | T/C | Known |
| 216 | 19 | 16559421 | rs58166386 | 4.42E-24/2.33E-10 | -0.040 (0.006) | A/G | Known |
| 217 | 19 | 18301979 | rs4808760 | 5.83E-25/2.17E-11 | 0.044 (0.007) | C/G | Known |
| 218 | 19 | 45143942 | rs7260482 | 1.43E-09/0.0008253 | -0.022 (0.007) | A/C | Known |
| 219 | 19 | 47638539 | rs11083862 | 2.44E-09/1.68E-08 | 0.033 (0.006) | A/T | Known |
| 220 | 19 | 49837246 | rs1465697 | 3.02E-18/5.12E-13 | 0.050 (0.007) | T/C | Known |
| 221 | 20 | 39968188 | rs6072343 | 2.69E-09/0.0111 | 0.022 (0.008) | A/G | Known |
| 222 | 20 | 42579051 | rs4812772 | 6.17E-09/0.003712 | -0.019 (0.006) | T/C | Known |
| 223 | 20 | 44734310 | rs6032662 | 5.28E-19/1.61E-14 | -0.051 (0.007) | T/C | Known |
| 224 | 20 | 47253487 | rs3935549 | 1.62E-08 | -0.033 (0.006) | T/C | Novel |
| 225 | 20 | 48422095 | rs6020055 | 5.12E-08/0.06806 | 0.026 (0.014) | A/G | Known |
| 226 | 20 | 52744437 | rs2585447 | 3.13E-10/0.005717 | -0.020 (0.007) | T/C | Known |
| 227 | 20 | 52789743 | rs2248137 | 1.92E-19/1.12E-10 | 0.039 (0.006) | C/G | Known |
| 228 | 20 | 62374441 | rs6742 | 4.11E-14/1.30E-07 | -0.047 (0.009) | T/C | Known |
| 229 | 21 | 34787312 | rs9808753 | 1.60E-09/0.001525 | -0.027 (0.008) | A/G | Known |
| 230 | 21 | 39864727 | rs2836438 | 2.62E-10/4.48E-06 | 0.043 (0.009) | A/G | Known |
| 231 | 22 | 22205353 | rs9610458 | 1.16E-19/1.17E-11 | 0.040 (0.006) | T/C | Known |
| 232 | 22 | 31622539 | rs4820955 | 1.06E-08 |  |  | Known |
| 233 | 22 | 37258986 | rs760517 | 5.22E-11/3.34E-06 | -0.028 (0.006) | T/C | Known |
| 234 | 22 | 37310954 | rs5756405 | 5.35E-11/3.41E-05 | 0.024 (0.006) | A/G | Known |
| 235 | 22 | 40291807 | rs137955 | 9.63E-09/0.0002301 | 0.022 (0.006) | T/C | Known |
| 236 | 22 | 50971266 | rs140522 | 1.31E-21/7.19E-12 | 0.043 (0.006) | T/C | Known |

**Table S3. SNPs that show some evidence of replication among AFR/AMR participants.**

|  | | | | | **AFR** | | | **AMR** | | | **EUR** | | |
| --- | --- | --- | --- | --- | --- | --- | --- | --- | --- | --- | --- | --- | --- |
| **SNP** | **Chr** | **Pos** | **A1** | **A2** | **EAF** | **Beta** | **P** | **EAF** | **Beta** | **P** | **EAF** | **Beta** | **P** |
| 1:6512547 | 1 | 6512547 | T | C | 0.160 | -0.241 | 9.52E-04 | 0.781 | -0.154 | 1.29E-01 | 0.795 | -0.034 | 8.91E-17 |
| 1:85764886 | 1 | 85764886 | A | G | 0.598 | -0.134 | 2.49E-02 | 0.656 | -0.022 | 7.86E-01 | 0.597 | -0.041 | 6.12E-12 |
| 1:92222089 | 1 | 92222089 | T | C | 0.162 | -0.226 | 4.12E-03 | 0.177 | 0.043 | 6.88E-01 | 0.154 | -0.051 | 3.68E-10 |
| 2:151644203 | 2 | 151644203 | T | C | 0.576 | -0.130 | 3.06E-02 | 0.775 | 0.091 | 3.12E-01 | 0.698 | -0.024 | 8.28E-09 |
| 2:191989356 | 2 | 191989356 | A | C | 0.539 | -0.172 | 3.39E-03 | 0.258 | 0.055 | 5.22E-01 | 0.342 | -0.029 | 2.40E-13 |
| 3:105455955 | 3 | 105455955 | T | C | 0.431 | -0.121 | 4.17E-02 | 0.406 | -0.034 | 6.74E-01 | 0.340 | -0.033 | 7.67E-08 |
| 3:141150990 | 3 | 141150990 | A | G | 0.092 | -0.239 | 8.22E-03 | 0.166 | -0.022 | 8.30E-01 | 0.311 | -0.020 | 1.30E-09 |
| 4:40307564 | 4 | 40307564 | T | C | 0.427 | -0.140 | 1.95E-02 | 0.708 | -0.118 | 1.55E-01 | 0.664 | -0.035 | 2.42E-08 |
| 5:133891282 | 5 | 133891282 | T | C | 0.355 | -0.120 | 4.65E-02 | 0.450 | -0.067 | 3.93E-01 | 0.508 | -0.028 | 1.91E-13 |
| 6:159465977 | 6 | 159465977 | T | C | 0.702 | -0.183 | 4.20E-03 | 0.555 | -0.030 | 7.06E-01 | 0.433 | -0.043 | 5.22E-13 |
| 7:56091706 | 7 | 56091706 | A | G | 0.576 | -0.119 | 4.70E-02 | 0.326 | 0.041 | 6.18E-01 | 0.273 | -0.038 | 5.30E-09 |
| 8:79417222 | 8 | 79417222 | A | G | 0.462 | 0.145 | 1.58E-02 | 0.716 | 0.137 | 1.15E-01 | 0.680 | -0.040 | 2.07E-10 |
| 11:118747813 | 11 | 118747813 | A | G | 0.070 | 0.188 | 3.66E-02 | 0.464 | -0.025 | 7.55E-01 | 0.453 | 0.033 | 1.40E-08 |
| 12:6440009 | 12 | 6440009 | T | C | 0.639 | -0.166 | 7.45E-03 | 0.719 | -0.056 | 4.92E-01 | 0.575 | -0.048 | 3.54E-16 |
| 16:11114512 | 16 | 11114512 | A | G | 0.232 | 0.179 | 5.96E-03 | 0.656 | -0.003 | 9.75E-01 | 0.589 | 0.063 | 2.90E-26 |
| 17:34842521 | 17 | 34842521 | A | G | 0.445 | -0.142 | 2.85E-02 | 0.438 | -0.085 | 3.09E-01 | 0.550 | -0.033 | 2.77E-08 |
| 18:56269737 | 18 | 56269737 | A | G | 0.506 | 0.130 | 2.77E-02 | 0.463 | 0.041 | 5.96E-01 | 0.519 | 0.015 | 3.90E-10 |
| 19:49837246 | 19 | 49837246 | T | C | 0.358 | 0.158 | 1.82E-02 | 0.246 | 0.007 | 9.41E-01 | 0.238 | 0.050 | 5.12E-13 |
| 1:2520527 | 1 | 2520527 | T | C | 0.200 | 0.026 | 7.01E-01 | 0.512 | 0.216 | 7.05E-03 | 0.667 | 0.049 | 2.50E-15 |
| 1:117090493 | 1 | 117090493 | T | C | 0.778 | -0.017 | 8.05E-01 | 0.363 | -0.185 | 3.31E-02 | 0.138 | -0.087 | 1.45E-24 |
| 2:68646536 | 2 | 68646536 | T | C | 0.385 | 0.062 | 3.00E-01 | 0.667 | 0.194 | 2.08E-02 | 0.550 | 0.029 | 5.08E-16 |
| 3:101749022 | 3 | 101749022 | T | C | 0.542 | 0.036 | 5.45E-01 | 0.683 | -0.191 | 2.07E-02 | 0.653 | -0.041 | 1.98E-11 |
| 3:159691112 | 3 | 159691112 | T | C | 0.666 | 0.021 | 7.43E-01 | 0.637 | 0.158 | 4.97E-02 | 0.541 | -0.042 | 8.46E-13 |
| 4:103911781 | 4 | 103911781 | A | G | 0.716 | -0.055 | 3.88E-01 | 0.553 | 0.166 | 3.51E-02 | 0.488 | 0.027 | 7.03E-16 |
| 10:8098719 | 10 | 8098719 | T | C | 0.523 | 0.004 | 9.46E-01 | 0.147 | -0.201 | 4.88E-02 | 0.164 | -0.035 | 7.16E-11 |
| 11:64095178 | 11 | 64095178 | T | C | 0.983 | 0.069 | 5.97E-01 | 0.847 | 0.261 | 1.07E-02 | 0.697 | -0.027 | 1.12E-11 |
| 14:88407917 | 14 | 88407917 | A | G | 0.181 | -0.004 | 9.49E-01 | 0.605 | 0.205 | 1.06E-02 | 0.512 | 0.041 | 4.08E-12 |
| 19:16559421 | 19 | 16559421 | A | G | 0.441 | -0.077 | 2.10E-01 | 0.627 | -0.163 | 4.92E-02 | 0.692 | -0.040 | 2.33E-10 |
| 20:39968188 | 20 | 39968188 | A | G | 0.067 | -0.020 | 8.46E-01 | 0.112 | 0.302 | 1.01E-02 | 0.139 | 0.022 | 2.69E-09 |

**Table S4: The shared effect of rs3184504 in seven autoimmune diseases.**

| **Trait** | **Tested Allele** | **Other Allele** | **Effect size** | | **P** |
| --- | --- | --- | --- | --- | --- |
| Celiac disease | T | C | 0.011 | 4.98E-10 | |
| IBD | T | C | 0.077 | 6.88E-10 | |
| MS | T | T | 4.550 | 5.37E-06 | |
| Psoriasis | T | T | 0.088 | 2.60E-08 | |
| SLE | T | C | 5.483 | 4.18E-08 | |
| T1D | T | C | 0.231 | 1.08E-60 | |
| Thyroiditis | T | C | 0.179 | 1.40E-25 | |

**Table S5. Genome-wide Association Study Summary Statistics used in MS genetic overlap analysis.**

| **Category** | **Trait** | **Abbreviation** | **Year** | **Ancestry** | **Assembly** | **PMID** |
| --- | --- | --- | --- | --- | --- | --- |
| **Neurodegenerative disease** | Alzheimer's disease | AD | 2022 | European | GRCh37/hg19 | 35379992 |
|  | Amyotrophic lateral sclerosis | ALS | 2021 | European | GRCh37/hg19 | 34873335 |
|  | Frontotemporal dementia | FTD | 2014 | European | GRCh37/hg19 | 24943344 |
|  | Parkinson's disease | PD | 2019 | European | GRCh37/hg19 | 31701892 |
| **Autoimmune/inflammatory diseases** | IgA nephropathy | IGA | 2021 | European | GRCh37/hg19 | 34594039 |
|  | Chronic obstructive pulmonary disease | COPD | 2021 | European | GRCh37/hg19 | 34737426 |
|  | Obesity | OB | 2021 | European | GRCh37/hg19 | 34737426 |
|  | Psoriasis | PS | 2022 | European | GRCh37/hg19 | 34927100 |
|  | Rheumatoid arthritis | RA | 2014 | European | GRCh37/hg19 | 24390342 |
|  | Systemic lupus erythematosus | SLE | 2018 | European | GRCh37/hg19 | 29848360 |
|  | Type 1 diabetes | T1D | 2021 | European | GRCh37/hg19 | 34012112 |
|  | Thyroiditis | TRD | 2021 | European | GRCh37/hg19 | 34594039 |
|  | Inflammatory bowel disease | IBD | 2017 | European | GRCh37/hg19 | 28067908 |
|  | Crohn's disease | CD | 2017 | European | GRCh37/hg19 | 28067908 |
|  | Celiac disease | CeD | 2021 | European | GRCh37/hg19 | 34278373 |
|  | Ulcerative colitis | UC | 2017 | European | GRCh37/hg19 | 28067908 |
| **Psychiatric disorders/trait** | Bipolar disorder | BIP | 2021 | European | GRCh37/hg19 | 34002096 |
|  | Major depressive disorder | MDD | 2019 | European | GRCh37/hg19 | 30718901 |
|  | Neuroticism | Neuro | 2018 | European | GRCh37/hg19 | 29942085 |
|  | Schizophrenia | SCZ | 2022 | European | GRCh37/hg19 | 35396580 |
| **BMI traits** | Type 2 diabetes | T2D | 2018 | European | GRCh37/hg19 | 30054458 |
|  | Body mass index | BMI | 2019 | European | GRCh37/hg19 | 30239722 |
|  | waist-to-hip ratio adjusted BMI | WHRadjBMI | 2019 | European | GRCh37/hg19 | 30239722 |

**Table S6. Genetic correlation between MS and other 24 traits.**

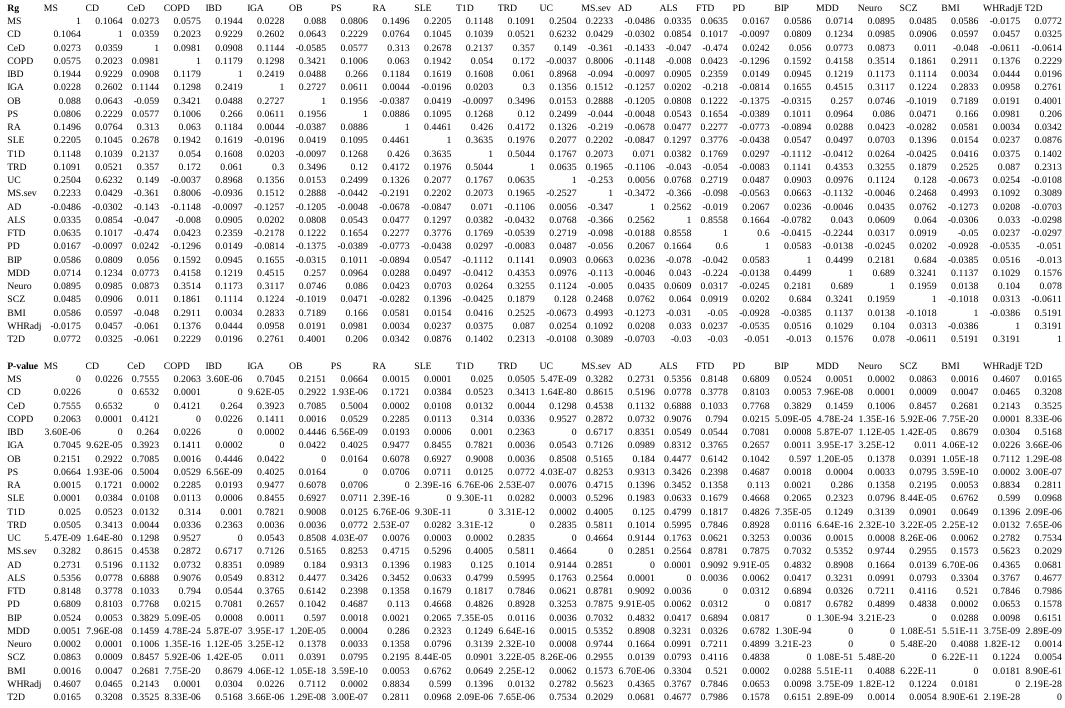

**Table S7. Colocalization between risk loci of MS and 20 cell type-level eQTL.**

PP.H4 values computed by coloc.abf are shown.

| **Trait** | **Cell type** | **Lead SNP** | **eGene** | **PP4** |
| --- | --- | --- | --- | --- |
| MS | Ast | rs10245867 | JAZF1 | 0.8724 |
| MS | Ast | rs12147246 | TRAF3 | 0.8961 |
| MS | Ast | rs12588969 | TRAF3 | 0.8961 |
| MS | Ast | rs13385171 | AC012370.2 | 0.9619 |
| MS | Ast | rs249677 | NDFIP1 | 0.9268 |
| MS | Ast | rs35218683 | IFITM3 | 0.9866 |
| MS | Ast | rs3809627 | KCTD13 | 0.8306 |
| MS | Ast | rs3923387 | PLEC | 0.9044 |
| MS | Ast | rs4796224 | MYO19 | 0.9082 |
| MS | Ast | rs56232455 | IFITM3 | 0.9866 |
| MS | Ast | rs570429157 | RRAS2 | 0.9748 |
| MS | Ast | rs61884005 | RRAS2 | 0.9744 |
| MS | Ast | rs701006 | AC025165.2 | 0.8402 |
| MS | BIN | rs11231749 | AP003774.1 | 0.8835 |
| MS | BIN | rs12434551 | ZFP36L1 | 0.9758 |
| MS | BIN | rs13218824 | AHI1 | 0.9870 |
| MS | BIN | rs1323292 | RGS1 | 0.9465 |
| MS | BIN | rs13414105 | LBH | 0.9730 |
| MS | BIN | rs140522 | TYMP | 0.9738 |
| MS | BIN | rs1465697 | CD37 | 0.9980 |
| MS | BIN | rs2317231 | FCRL3 | 0.8378 |
| MS | BIN | rs2327586 | AHI1 | 0.9870 |
| MS | BIN | rs28703878 | ZC2HC1A | 0.8853 |
| MS | BIN | rs35218683 | RP11-326C3.11 | 0.9606 |
| MS | BIN | rs4896153 | AHI1 | 0.9870 |
| MS | BIN | rs4952115 | LBH | 0.9730 |
| MS | BIN | rs56232455 | RP11-326C3.11 | 0.9606 |
| MS | BIN | rs76892387 | AHI1 | 0.9870 |
| MS | BIN | rs77191363 | FCRL3 | 0.8378 |
| MS | BIN | rs7977720 | CLEC2D | 0.9950 |
| MS | BMem | rs1076928 | ETV7 | 0.8698 |
| MS | BMem | rs11231749 | AP003774.1 | 0.8617 |
| MS | BMem | rs12365699 | DDX6 | 0.8639 |
| MS | BMem | rs12434551 | ZFP36L1 | 0.9850 |
| MS | BMem | rs13218824 | AHI1 | 0.9878 |
| MS | BMem | rs140522 | TYMP | 0.9894 |
| MS | BMem | rs1465697 | CD37 | 0.9893 |
| MS | BMem | rs149114341 | DDX6 | 0.8639 |
| MS | BMem | rs2327586 | AHI1 | 0.9878 |
| MS | BMem | rs28703878 | ZC2HC1A | 0.8677 |
| MS | BMem | rs34026809 | DDX6 | 0.8640 |
| MS | BMem | rs35218683 | IFITM2 | 0.9978 |
| MS | BMem | rs4896153 | AHI1 | 0.9878 |
| MS | BMem | rs56232455 | IFITM2 | 0.9978 |
| MS | BMem | rs6589706 | DDX6 | 0.8639 |
| MS | BMem | rs76892387 | AHI1 | 0.9878 |
| MS | BMem | rs7977720 | CLEC2D | 0.9390 |
| MS | CD4ET | rs11230581 | CD6 | 0.9927 |
| MS | CD4ET | rs11231749 | AP003774.1 | 0.8887 |
| MS | CD4ET | rs13218824 | AHI1 | 0.9875 |
| MS | CD4ET | rs13414105 | LBH | 0.8622 |
| MS | CD4ET | rs1465697 | CD37 | 0.9847 |
| MS | CD4ET | rs2327586 | AHI1 | 0.9875 |
| MS | CD4ET | rs28703878 | ZC2HC1A | 0.8674 |
| MS | CD4ET | rs4409785 | SESN3 | 0.8447 |
| MS | CD4ET | rs4896153 | AHI1 | 0.9875 |
| MS | CD4ET | rs4939490 | CD6 | 0.9927 |
| MS | CD4ET | rs4952115 | LBH | 0.8622 |
| MS | CD4ET | rs531612 | BANF1 | 0.8606 |
| MS | CD4ET | rs56095240 | SESN3 | 0.8461 |
| MS | CD4ET | rs75064517 | CD6 | 0.9927 |
| MS | CD4ET | rs76892387 | AHI1 | 0.9875 |
| MS | CD4ET | rs7731626 | ANKRD55 | 0.9994 |
| MS | CD4NC | rs10245867 | JAZF1 | 0.8631 |
| MS | CD4NC | rs1076928 | ETV7 | 0.9394 |
| MS | CD4NC | rs11230581 | CD6 | 0.9971 |
| MS | CD4NC | rs11231749 | AP003774.1 | 0.8771 |
| MS | CD4NC | rs11542663 | ASF1A | 0.9188 |
| MS | CD4NC | rs12365699 | DDX6 | 0.8475 |
| MS | CD4NC | rs12434551 | ZFP36L1 | 0.9658 |
| MS | CD4NC | rs12622670 | PLEK | 0.8929 |
| MS | CD4NC | rs13218824 | AHI1 | 0.9881 |
| MS | CD4NC | rs1465697 | CD37 | 0.9747 |
| MS | CD4NC | rs149114341 | DDX6 | 0.8475 |
| MS | CD4NC | rs2327586 | AHI1 | 0.9881 |
| MS | CD4NC | rs28703878 | ZC2HC1A | 0.8075 |
| MS | CD4NC | rs34026809 | DDX6 | 0.8475 |
| MS | CD4NC | rs375915427 | MMEL1 | 0.8450 |
| MS | CD4NC | rs3923387 | PLEC | 0.8710 |
| MS | CD4NC | rs4409785 | SESN3 | 0.9982 |
| MS | CD4NC | rs4796224 | GGNBP2 | 0.9024 |
| MS | CD4NC | rs4896153 | AHI1 | 0.9881 |
| MS | CD4NC | rs4939490 | CD6 | 0.9971 |
| MS | CD4NC | rs531612 | BANF1 | 0.9725 |
| MS | CD4NC | rs531612 | DRAP1 | 0.9618 |
| MS | CD4NC | rs56095240 | SESN3 | 0.9983 |
| MS | CD4NC | rs6589706 | DDX6 | 0.8475 |
| MS | CD4NC | rs6670198 | MMEL1 | 0.8450 |
| MS | CD4NC | rs75064517 | CD6 | 0.9971 |
| MS | CD4NC | rs75191738 | TMEM244 | 0.8248 |
| MS | CD4NC | rs76892387 | AHI1 | 0.9881 |
| MS | CD4NC | rs7731626 | ANKRD55 | 0.9994 |
| MS | CD4NC | rs7731626 | IL6ST | 0.9994 |
| MS | CD4NC | rs77654077 | GPR18 | 0.9274 |
| MS | CD4NC | rs7975763 | MPHOSPH9 | 0.8617 |
| MS | CD4NC | rs8062446 | NLRC5 | 0.9991 |
| MS | CD4NC | rs883871 | NR1D1 | 0.9825 |
| MS | CD4NC | rs883871 | THRA | 0.9942 |
| MS | CD4NC | rs9909593 | NR1D1 | 0.9826 |
| MS | CD4NC | rs9909593 | THRA | 0.9943 |
| MS | CD8ET | rs1076928 | ETV7 | 0.9099 |
| MS | CD8ET | rs10951042 | CARD11 | 0.9576 |
| MS | CD8ET | rs11231749 | AP003774.1 | 0.8902 |
| MS | CD8ET | rs11231749 | PRDX5 | 0.9072 |
| MS | CD8ET | rs11542663 | ASF1A | 0.8331 |
| MS | CD8ET | rs12622670 | PLEK | 0.9293 |
| MS | CD8ET | rs13218824 | AHI1 | 0.9895 |
| MS | CD8ET | rs137955 | ATF4 | 0.8588 |
| MS | CD8ET | rs1465697 | CD37 | 0.9690 |
| MS | CD8ET | rs2327586 | AHI1 | 0.9895 |
| MS | CD8ET | rs4796224 | GGNBP2 | 0.9255 |
| MS | CD8ET | rs4896153 | AHI1 | 0.9895 |
| MS | CD8ET | rs62013236 | CTSH | 0.9317 |
| MS | CD8ET | rs75191738 | TMEM244 | 0.8583 |
| MS | CD8ET | rs76892387 | AHI1 | 0.9895 |
| MS | CD8ET | rs9900529 | LLGL2 | 0.8878 |
| MS | CD8NC | rs1076928 | ETV7 | 0.9149 |
| MS | CD8NC | rs11230581 | CD6 | 0.9406 |
| MS | CD8NC | rs11231749 | AP003774.1 | 0.8844 |
| MS | CD8NC | rs12365699 | DDX6 | 0.8155 |
| MS | CD8NC | rs12434551 | ZFP36L1 | 0.8776 |
| MS | CD8NC | rs12622670 | PLEK | 0.8483 |
| MS | CD8NC | rs13218824 | AHI1 | 0.9875 |
| MS | CD8NC | rs1465697 | CD37 | 0.8701 |
| MS | CD8NC | rs149114341 | DDX6 | 0.8155 |
| MS | CD8NC | rs2327586 | AHI1 | 0.9875 |
| MS | CD8NC | rs34026809 | DDX6 | 0.8155 |
| MS | CD8NC | rs4409785 | SESN3 | 0.9980 |
| MS | CD8NC | rs4896153 | AHI1 | 0.9875 |
| MS | CD8NC | rs4939490 | CD6 | 0.9406 |
| MS | CD8NC | rs531612 | BANF1 | 0.9399 |
| MS | CD8NC | rs56095240 | SESN3 | 0.9980 |
| MS | CD8NC | rs6589706 | DDX6 | 0.8155 |
| MS | CD8NC | rs67111717 | RGS14 | 0.9872 |
| MS | CD8NC | rs75064517 | CD6 | 0.9406 |
| MS | CD8NC | rs76892387 | AHI1 | 0.9875 |
| MS | CD8NC | rs7731626 | ANKRD55 | 0.9994 |
| MS | CD8NC | rs7731626 | IL6ST | 0.9682 |
| MS | CD8NC | rs77654077 | GPR18 | 0.9193 |
| MS | CD8NC | rs7977720 | CLECL1 | 0.8495 |
| MS | CD8NC | rs883871 | THRA | 0.8428 |
| MS | CD8NC | rs9909593 | THRA | 0.8441 |
| MS | CD8S100B | rs11231749 | AP003774.1 | 0.8585 |
| MS | CD8S100B | rs12622670 | PLEK | 0.8838 |
| MS | CD8S100B | rs13218824 | AHI1 | 0.9845 |
| MS | CD8S100B | rs13414105 | LBH | 0.8847 |
| MS | CD8S100B | rs2327586 | AHI1 | 0.9845 |
| MS | CD8S100B | rs4896153 | AHI1 | 0.9845 |
| MS | CD8S100B | rs4952115 | LBH | 0.8847 |
| MS | CD8S100B | rs76892387 | AHI1 | 0.9845 |
| MS | DC | rs1323292 | RGS1 | 0.9767 |
| MS | DC | rs140522 | TYMP | 0.9989 |
| MS | DC | rs35218683 | IFITM3 | 0.9983 |
| MS | DC | rs56232455 | IFITM3 | 0.9983 |
| MS | DC | rs7975763 | ARL6IP4 | 0.8185 |
| MS | Exc | rs10245867 | JAZF1 | 0.9440 |
| MS | Exc | rs11542663 | AL137009.1 | 0.9844 |
| MS | Exc | rs13218824 | LINC00271 | 0.8935 |
| MS | Exc | rs2327586 | LINC00271 | 0.8935 |
| MS | Exc | rs28703878 | PKIA-AS1 | 0.8655 |
| MS | Exc | rs3923387 | PLEC | 0.8893 |
| MS | Exc | rs4796224 | MYO19 | 0.9953 |
| MS | Exc | rs4896153 | LINC00271 | 0.8935 |
| MS | Exc | rs6032662 | SLC12A5 | 0.9305 |
| MS | Exc | rs6072343 | ZHX3 | 0.9045 |
| MS | Exc | rs6738544 | STAT4 | 0.9167 |
| MS | Exc | rs6742 | ZBTB46 | 0.9815 |
| MS | Exc | rs701006 | AGAP2 | 0.8126 |
| MS | Exc | rs76892387 | LINC00271 | 0.8935 |
| MS | Exc | rs9610458 | AC245452.1 | 0.9165 |
| MS | Inh | rs1026916 | STAT3 | 0.9730 |
| MS | Inh | rs10936602 | LRRC34 | 0.9374 |
| MS | Inh | rs11542663 | AL137009.1 | 0.9876 |
| MS | Inh | rs12147246 | TRAF3 | 0.8416 |
| MS | Inh | rs12588969 | TRAF3 | 0.8416 |
| MS | Inh | rs13218824 | LINC00271 | 0.9804 |
| MS | Inh | rs2289746 | CBLB | 0.9824 |
| MS | Inh | rs2327586 | LINC00271 | 0.9804 |
| MS | Inh | rs28703878 | IL7 | 0.8747 |
| MS | Inh | rs28703878 | PKIA-AS1 | 0.8691 |
| MS | Inh | rs3923387 | PLEC | 0.9053 |
| MS | Inh | rs4796224 | DHRS11 | 0.8638 |
| MS | Inh | rs4796224 | MYO19 | 0.9316 |
| MS | Inh | rs4896153 | LINC00271 | 0.9804 |
| MS | Inh | rs58905292 | STAT3 | 0.9730 |
| MS | Inh | rs6032662 | SLC12A5 | 0.9314 |
| MS | Inh | rs67934705 | FUCA1 | 0.9737 |
| MS | Inh | rs76892387 | LINC00271 | 0.9804 |
| MS | Inh | rs9308424 | FLVCR1 | 0.9287 |
| MS | Inh | rs9900529 | AC011933.4 | 0.9254 |
| MS | Mic | rs10245867 | JAZF1 | 0.9147 |
| MS | Mic | rs11231749 | VEGFB | 0.9055 |
| MS | Mic | rs13218824 | AHI1 | 0.9753 |
| MS | Mic | rs2327586 | AHI1 | 0.9753 |
| MS | Mic | rs28703878 | ZC2HC1A | 0.8733 |
| MS | Mic | rs3923387 | PLEC | 0.9622 |
| MS | Mic | rs4896153 | AHI1 | 0.9753 |
| MS | Mic | rs76892387 | AHI1 | 0.9753 |
| MS | Mic | rs9900529 | AC011933.4 | 0.8253 |
| MS | MonoC | rs11125803 | ADCY3 | 0.8111 |
| MS | MonoC | rs1323292 | RGS1 | 0.9828 |
| MS | MonoC | rs140522 | TYMP | 0.9986 |
| MS | MonoC | rs35218683 | IFITM3 | 0.9984 |
| MS | MonoC | rs56232455 | IFITM3 | 0.9984 |
| MS | MonoNC | rs1323292 | RGS1 | 0.9611 |
| MS | MonoNC | rs140522 | TYMP | 0.9988 |
| MS | MonoNC | rs35218683 | IFITM3 | 0.9984 |
| MS | MonoNC | rs56232455 | IFITM3 | 0.9984 |
| MS | MonoNC | rs6911131 | PEX3 | 0.9351 |
| MS | MonoNC | rs9308424 | BATF3 | 0.9960 |
| MS | NKR | rs10271373 | ZC3HAV1 | 0.8839 |
| MS | NKR | rs35218683 | IFITM3 | 0.9984 |
| MS | NKR | rs56232455 | IFITM3 | 0.9984 |
| MS | NK | rs1076928 | ETV7 | 0.9141 |
| MS | NK | rs11231749 | AP003774.1 | 0.8341 |
| MS | NK | rs11231749 | PRDX5 | 0.8829 |
| MS | NK | rs11231749 | TRMT112 | 0.9080 |
| MS | NK | rs11578655 | AC093157.1 | 0.9198 |
| MS | NK | rs12047318 | AC093157.1 | 0.9198 |
| MS | NK | rs12622670 | PLEK | 0.9015 |
| MS | NK | rs13218824 | AHI1 | 0.9882 |
| MS | NK | rs1399180 | GATA3 | 0.9350 |
| MS | NK | rs142860878 | AC093157.1 | 0.9198 |
| MS | NK | rs1465697 | CD37 | 0.9892 |
| MS | NK | rs147885102 | AC093157.1 | 0.9198 |
| MS | NK | rs2327586 | AHI1 | 0.9882 |
| MS | NK | rs34723276 | AC093157.1 | 0.9198 |
| MS | NK | rs35218683 | IFITM3 | 0.9984 |
| MS | NK | rs35218683 | RP11-326C3.15 | 0.9926 |
| MS | NK | rs4896153 | AHI1 | 0.9882 |
| MS | NK | rs56232455 | IFITM3 | 0.9984 |
| MS | NK | rs56232455 | RP11-326C3.15 | 0.9926 |
| MS | NK | rs76892387 | AHI1 | 0.9882 |
| MS | Oli | rs137955 | SYNGR1 | 0.8477 |
| MS | Oli | rs483180 | PHGDH | 0.8495 |
| MS | Oli | rs9900529 | AC011933.4 | 0.8794 |
| MS | OPC | rs10245867 | JAZF1 | 0.8883 |
| MS | OPC | rs11542663 | AL137009.1 | 0.9845 |

**Table S8: Validation of COLOC results using MIT Mathy’s snucRNAseq data and in-house multiome data.**

| **Cell type** | **SNP** | **Gene** | **Kellis (P-value)** | **Multiome (P-value)** | **Meta-analysis (P-value)** |
| --- | --- | --- | --- | --- | --- |
| Ast | rs10245867 | JAZF1 | 8.18E-02 | 7.51E-01 | 8.68E-02 |
| Ast | rs12147246 | TRAF3 | 3.88E-04 | 7.70E-05 | 1.69E-06 |
| Ast | rs12588969 | TRAF3 | 5.47E-01 | 1.21E-01 | 5.19E-02 |
| Ast | rs13385171 | AC012370.2 | NA | NA | NA |
| Ast | rs249677 | NDFIP1 | 1.45E-02 | 3.50E-03 | 2.48E-03 |
| Ast | rs35218683 | IFITM3 | 9.61E-08 | 6.55E-07 | 2.30E-05 |
| Ast | rs3809627 | KCTD13 | 9.64E-02 | 4.46E-03 | 9.63E-02 |
| Ast | rs3923387 | PLEC | 2.64E-05 | 8.68E-05 | 7.74E-05 |
| Ast | rs4796224 | MYO19 | 1.20E-03 | 1.86E-02 | 9.60E-07 |
| Ast | rs56232455 | IFITM3 | 5.53E-06 | 4.56E-05 | 1.35E-04 |
| Ast | rs570429157 | RRAS2 | NA | NA | NA |
| Ast | rs61884005 | RRAS2 | 1.59E-01 | 2.17E-01 | 3.70E-02 |
| Ast | rs701006 | AC025165.2 | NA | NA | NA |
| Exc | rs10245867 | JAZF1 | 7.44E-05 | 3.87E-03 | 1.83E-04 |
| Exc | rs11542663 | AL137009.1 | NA | 2.22E-12 | NA |
| Exc | rs13218824 | LINC00271 | NA | NA | NA |
| Exc | rs2327586 | LINC00271 | NA | 1.29E-01 | NA |
| Exc | rs28703878 | PKIA-AS1 | NA | 7.52E-02 | NA |
| Exc | rs3923387 | PLEC | 9.49E-18 | 2.76E-14 | 1.46E-16 |
| Exc | rs4796224 | MYO19 | 1.26E-07 | 8.46E-03 | 3.29E-05 |
| Exc | rs4896153 | LINC00271 | NA | 6.05E-01 | NA |
| Exc | rs6032662 | SLC12A5 | 5.71E-08 | 8.66E-10 | 5.80E-07 |
| Exc | rs6072343 | ZHX3 | 8.03E-03 | 5.31E-01 | 3.20E-01 |
| Exc | rs6738544 | STAT4 | 2.02E-05 | 2.05E-04 | 4.14E-03 |
| Exc | rs6742 | ZBTB46 | 7.38E-01 | 4.59E-01 | 9.71E-02 |
| Exc | rs701006 | AGAP2 | 9.18E-02 | 4.89E-01 | 4.19E-01 |
| Exc | rs76892387 | LINC00271 | NA | NA | NA |
| Exc | rs9610458 | AC245452.1 | NA | NA | NA |
| Inh | rs1026916 | STAT3 | 6.45E-05 | 1.82E-03 | 5.60E-04 |
| Inh | rs10936602 | LRRC34 | 2.30E-02 | 3.79E-03 | 3.23E-03 |
| Inh | rs11542663 | AL137009.1 | NA | 1.18E-11 | NA |
| Inh | rs12147246 | TRAF3 | 6.66E-10 | 5.54E-07 | 6.78E-08 |
| Inh | rs12588969 | TRAF3 | 2.43E-02 | 1.85E-02 | 5.42E-03 |
| Inh | rs13218824 | LINC00271 | NA | NA | NA |
| Inh | rs2289746 | CBLB | NA | NA | NA |
| Inh | rs2327586 | LINC00271 | NA | 3.98E-01 | NA |
| Inh | rs28703878 | IL7 | 9.25E-01 | 1.90E-03 | 4.68E-01 |
| Inh | rs28703878 | PKIA-AS1 | NA | 5.45E-02 | NA |
| Inh | rs3923387 | PLEC | 2.06E-16 | 6.54E-09 | 4.81E-13 |
| Inh | rs4796224 | DHRS11 | 1.06E-02 | 4.77E-02 | 6.38E-03 |
| Inh | rs4796224 | MYO19 | 1.37E-02 | 3.36E-01 | 1.34E-05 |
| Inh | rs4896153 | LINC00271 | NA | 1.72E-04 | NA |
| Inh | rs58905292 | STAT3 | 3.77E-03 | 5.20E-01 | 2.29E-01 |
| Inh | rs6032662 | SLC12A5 | 1.27E-09 | 8.11E-11 | 2.74E-09 |
| Inh | rs67934705 | FUCA1 | 1.04E-01 | 8.16E-01 | 6.36E-02 |
| Inh | rs76892387 | LINC00271 | NA | NA | NA |
| Inh | rs9308424 | FLVCR1 | 9.56E-01 | 9.88E-01 | 6.95E-01 |
| Inh | rs9900529 | AC011933.4 | NA | NA | NA |
| Mic | rs10245867 | JAZF1 | 2.69E-13 | 2.95E-13 | 4.48E-16 |
| Mic | rs11231749 | VEGFB | 1.01E-02 | NA | NA |
| Mic | rs13218824 | AHI1 | NA | NA | NA |
| Mic | rs2327586 | AHI1 | 9.25E-01 | 6.17E-01 | 1.40E-01 |
| Mic | rs28703878 | ZC2HC1A | 3.04E-01 | 7.17E-03 | 1.54E-01 |
| Mic | rs3923387 | PLEC | 5.02E-15 | NA | NA |
| Mic | rs4896153 | AHI1 | 7.49E-10 | 2.32E-06 | 1.24E-09 |
| Mic | rs76892387 | AHI1 | NA | NA | NA |
| Mic | rs9900529 | AC011933.4 | NA | NA | NA |
| Oli | rs137955 | SYNGR1 | 7.56E-01 | 2.64E-01 | 6.69E-02 |
| Oli | rs483180 | PHGDH | 4.65E-04 | 9.94E-04 | 1.90E-02 |
| Oli | rs9900529 | AC011933.4 | NA | NA | NA |

**Table S9. Descriptive statistics of the new MS GWAS data of the discovery phase.**

| phi | Cases/controls | OR 95% CI, P | AUC (Crude) | Variance explained |
| --- | --- | --- | --- | --- |
| E+00 | 615/53250 | 1.59 (1.47-1.72), P=2.34E-30 | 0.6897 (0.6073) | 0.0203 |
| E-01 |  | 1.61 (1.48-1.74), P=8.86 E-32 | 0.6915 (0.6113) | 0.0213 |
| E-02 |  | 1.61 (1.48-1.74), P=6.91E-32 | 0.694 (0.6153) | 0.0214 |
| E-04 |  | 1.62 (1.49-1.75), P=5.33E-32 | 0.7019 (0.6231) | 0.0217 |
| E-06 |  | 1.44 (1.33-1.56), P=4.86E-19 | 0.6867(0.5923) | 0.0125 |
| E-08 |  | 1.24 (1.14-1.34), P=1.49E-07 | 0.6673(0.5524) | 0.0043 |
